# Metataxonomic analysis demonstrates a shift in duodenal microbiota in South African patients with obstructive jaundice: A pilot study

**DOI:** 10.1101/2023.05.15.23289977

**Authors:** Benjamin Hart, Jasmin Patel, Pieter De Maayer, Ekene Emmanuel Nweke, Damon Bizos

## Abstract

The human gastrointestinal tract (GIT) is home to an abundance of diverse microorganisms, and the balance of this microbiome plays a vital role in maintaining a healthy GIT. The obstruction of the flow of bile into the duodenum, resulting in obstructive jaundice (OJ), has a major impact on the health of the affected individual. This study sought to identify changes in the duodenal microbiota in South African patients with OJ compared to those without this disorder. Mucosal biopsies were taken from the duodenum of nineteen jaundiced patients undergoing endoscopic retrograde cholangiopancreatography (ERCP) and nineteen control participants (non-jaundiced patients) undergoing gastroscopy. DNA extracted from the samples was subjected to 16S rRNA amplicon sequencing using the Ion S5 TM sequencing platform. Diversity metrics and statistical correlation analyses with the clinical data were performed to compare duodenal microbial communities in both groups. Differences in the mean distribution of the microbial communities in the jaundiced and non-jaundiced samples were observed; however, this difference did not reach statistical significance. Of note, there was a statistically significant difference between the mean distributions of bacteria comparing jaundiced patients with cholangitis to those without. On further subset analysis, a significant difference was observed between patients with benign (Cholelithiasis) and malignant disease, namely head of pancreas (HOP) mass (p-values of 0.01). Beta diversity analyses further revealed a significant difference between patients with stone and non-stone related disease when factoring in the Campylobacter-Like Organisms (CLO) test status (*p*=0.048). This study demonstrated a shift in the microbiota in jaundiced patients, especially considering some underlying conditions of the upper GI tract. Future studies should aim to verify these findings in a larger cohort.

## INTRODUCTION

Disorders of the biliary tract, such as cholelithiasis (gallstone disease) and malignant obstruction may result in decreased bile flow through the extrahepatic bile ducts into the gastrointestinal tract (GIT). These disorders affect a significant proportion of the world’s population [1]. Studies have shown that in the United States, 10-15% of adults have cholelithiasis and 1 million newly diagnosed cases are recorded each year [1]. In South Africa, there has been a dramatic rise in the rates of cholecystectomies as a result of cholelithiasis and its complications [2]. This predisposes a significant proportion of the population to complications associated with gallstone disease, including bile duct obstruction. In the United States, the incidence of bile duct obstruction is approximately 5 cases per 1,000 [3]. The obstruction of bile flow results in bile stasis, allowing microorganisms present within the duodenum, to migrate into and colonize the bile ducts. When the biliary system, which under normal circumstances is sterile, is colonized by bacteria, this can lead to the development of acute cholangitis, a life-threatening infection. Cholangitis requires an urgent procedure such as endoscopic retrograde pancreaticoduodenoscopy (ERCP) to allow drainage of the biliary system. This drainage allows the restoration of normal bile flow which allows the resolution of this disorder [29].

It is widely accepted that a healthy gut microbiome is essential for the overall well-being of the host [4]. The gut microbiota develops from a relatively simple community of organisms at birth, into a diverse pool [5]. With time, the host-bacterial relationship becomes mutually beneficial, establishing a symbiotic relationship. Broadly speaking, the gut microbiome plays a pivotal part in the metabolism, nutrition and immune functioning of the host [5]. It helps to prevent the colonization of the GIT with potentially pathogenic organisms and their subsequent overgrowth [6]. The GIT is home to an abundance and a wide diversity of microorganisms, with an excess of 1 x 10^10^ organisms inhabiting the GIT of a healthy person [5]. These microorganisms can be broadly classified into 5 predominant phyla, namely the Bacillota, Bacteroidota, Actinomycetota, Pseudomonadota and Verrucimicrobiota [7]. Members of the phyla Bacillota and Bacteroidota make up ∼98% of the overall gut microbiota and as such can be used as indicators or biomarkers of a healthy GIT [7,8]. Compared to the lower GIT, the microbiota of the proximal GIT is relatively understudied. A study observed significant differences in the duodenal microbiota compared to that of the rectum, with the duodenum containing a greater diversity of organisms, with the predominant genera being *Prevotella* and *Acinetobacter* from the phylum Bacteroidota and Pseudomonadota, respectively [9].

The small intestine plays a pivotal role in nutrient digestion and absorption. The duodenum is an important region of the small bowel and is responsible for the mixing of gastric chyme, pancreatic enzymes and bile products from the hepatobiliary tract. The process of bile synthesis, transport, secretion, reabsorption and elimination occurs in the liver and is pivotal to the normal functioning of the body [3]. The relationship between the GIT microbiota and bile is co-dependent with each one relying on the other to maintain homeostasis. Bile acids have important antimicrobial functions, preventing bacterial overgrowth and helping to maintain the homeostasis of the gut microbiome [10]. In turn, a healthy gut microbiota contributes in the bio-transformation of bile acids which helps to maintain the normal cycle of bile acid production, secretion and reabsorption [11]. The microbiota mediates the biotransformation of bile acids, this subsequently results in the activation of bile acid receptors, for example, nuclear receptor farnesoid X which governs bile, glucose and lipid metabolism [12].

In this pilot study, mucosal biopsies of the duodenum were analysed to explore the differences between the microbiomes of patients with obstructive jaundiced group compared to a control group using 16S rRNA amplicon sequencing analysis. A shift in duodenal microbiota in jaundiced patients compared to the control was observed. Furthermore, the study demonstrated a difference in the mean distribution of the microbial community in jaundiced patients with cholangitis versus those without cholangitis.

## MATERIALS AND METHODS

### Study site and population

This study was conducted at the Endoscopy suite of Charlette Maxeke Johannesburg Academic Hospital (CMJAH), Johannesburg, Gauteng, South Africa. Patients were prospectively recruited between June 2021 to December 2021. Ethical approval was obtained from the Human Research Ethics Committee of the University of the Witwatersrand, Johannesburg, South Africa (Ethics number: M200908).

A pilot cohort of 38 individuals was recruited including nineteen patients with jaundice and nineteen without jaundice (control group). Patients were included as part of the jaundice group if they were at least 18 years old, diagnosed clinically with obstructive jaundice having a proven common bile duct (CBD) dilation using ultrasound or computerized tomography (CT) scan. The control group included individuals who were at least 18 years old, with no obstructive jaundice. For both groups, individuals who had any endoscopy or antibiotics in the last 3 months were excluded. Relevant demographic and clinicopathological parameters such as age, gender and any underlying diseases were collected from each participant. All participants gave written informed consent before enrolment into the study.

### Sample collection and CLO testing

Mucosal biopsies were obtained by endoscopic retrograde cholangiopancreatography (ERCP) and gastroscopy from each of the jaundiced and control participants, respectively. The presence or absence of *Helicobacter pylori* in mucosal biopsies of the gastric antrum was confirmed using standard *Campylobacter-*like organism (CLO) testing (Kim et al., 2000). Additionally, mucosal biopsies were taken from the second part (descending portion) of the duodenum and placed in a DNA/RNA shield solution (Zymo Research, Irvine, CA 92614, U.S.A.). Samples were stored at 4 °C for 24 hrs before being transferred to a -20 °C freezer.

### DNA extraction, quantification and quality control

Genomic DNA extraction from the collected tissue was performed using the QIAamp DNA mini kit (Qiagen, Hilden, Germany), as per the manufacturer’s protocol. The extracted metagenomic DNA (mDNA) were quantified using the Qubit 1x dsDNA HS assay kit (ThermoFisher Scientific, Waltham, MA, U.S.A) on the Qubit 4.0 Fluorometer as per the manufacturer’s protocol. The purity of the samples was assessed using the NanoDrop ND-1000 (ThermoFischer Scientific). The Genomic DNA (gDNA) Reagent Kit and the DNA Extended Range Chip (PerkinElmer, Waltham, MA, USA) were used to determine the genomic quality scores (GQS) using the LabChip GX Touch 24 Nucleic Acid Analyzer, using as per the manufacturer’s protocol.

### 16S rRNA amplification, library preparation and sequencing

The Ion 16STM Metagenomics Kit (ThermoFisher Scientific, USA) was used to amplify hypervariable regions from polybacterial samples according to the manufacturer’s protocol (MAN0010799 REV C.0). Amplification of target regions was performed from 2µl mDNA using the SimpliAmp Thermal Cycler (ThermoFisher Scientific, USA) for 25 cycles. Verification of the amplified products was performed utilizing the HT DNA NGS 3K reagent kit and X-mark chip on the PerkinElmer LabChip GXII Touch (Perkin Elmer, USA) as per the manufacturer’s protocol. Afterwards, polymerase chain reaction products from the primer pools were combined for each sample, purified with AgencourtTM AMPureTM XP (Beckman Coulter, Brea, CA, USA) reagent and eluted in 15 µl nuclease-free water. The quantification of purified amplicons was done using the Qubit 1x dsDNA HS assay kit on the Qubit 4.0 Fluorometer according to the manufacturer’s protocol.

Library preparation was conducted from 100 ng pooled amplicons for each sample using the Ion Plus Fragment Library Kit as per the manufacturer’s protocol. Purification of the end-repaired products was performed using the AgencourtTM AMPureTM XP (Beckman Coulter) reagent. The end-repaired product was ligated to 2ul IonCodeTM Barcode Adapters (ThermoFisher Scientific, USA). The adapter-ligated, barcoded libraries were purified with 1.4x Agencourt AMPureTM XP reagent (Beckman Coulter) and quantified using the Ion Universal Library Quantitation Kit according to the manufacturer’s protocol. qPCR amplification was then conducted using the StepOnePlusTM Real-time PCR system (ThermoFisher Scientific). Assessment of the library fragment size distributions was conducted on the LabChip® GXII Touch (PerkinElmer, Waltham, MA, USA), using the X-mark chip and HT DNA NGS 3K reagent kit as per the manufacturer’s protocol. The libraries were diluted to a concentration of 15 pM. The Ion 510TM, Ion 520TM & Ion 530TM Chef Kit (ThermoFisher Scientific, USA) were used to combine equimolar amounts for template preparation of the diluted, barcoded, 16S libraries. Summarily, the pooled library (25 µl) was loaded on the Ion Chef liquid handler. Enriched, template-positive ion sphere particles were transferred onto an Ion 530TM Chip (ThermoFisher Scientific, USA). Massively parallel sequencing was conducted using the Ion S5TM sequencing (ThermoFisher Scientific, USA). Sequencing Re-agents according to the manufacturer’s protocol (MAN0016854, REV.F.0) on the Ion S5TM Prime. Using default analysis parameters in the Torrent Suite Version 5.16.0 Software, flow space calibration and BaseCaller analyses were conducted. The subsequent run data was uploaded to the relevant Ion Reporter cloud account. The Ion Re-porter Metagenomics 16S w1.1 workflow was employed for OTU binning (99.0% species and 97.0% genus cut-off threshold) and taxonomic classification against the curated Greengenes V13.5 and MicroSEQ® 16S v2013.1 reference libraries [13]

### Microbial diversity analyses

The processed OTU and taxonomy tables were analysed using R v 3.6.1 (R Foundation for Statistical Computing; http://www.R-project.org) and RStudio (RStudio Team, 2014). Singleton sequences were first removed after which the dataset was rarefied to standardise the sample sizes [14]. The bacterial 16S rRNA reads were rarefied to 37,953 reads to reflect the lowest number of sequences after all low abundance counts from the samples were removed. The multivariant test using Dirichlet-Multinomial distribution was employed to test for differences in the mean distribution of bacterial taxa at the phylum level using the function Xdc.sevsample from HMP [15]. Bacterial α-diversity was then estimated using the observed OTU richness and the Shan-non-Weaver diversity index. The data distribution of the α-diversity indices was determined using the Shapiro-Wilk test for normality. Analysis of Variance (ANOVA) and Tukey’s Honest Significant Difference test (Tukey’s HSD) were performed to test the relationship between the α-diversity indices and the various clinical diagnosis, gender, and CLO test results of the patients (categorical variables). The general linear model using the function glm from STAT (R Core Team, 2021) was used to test the relationship between the patient’s age (continuous variable) with the α-diversity indices. Bacterial β-diversity was analysed based on the relative abundance of OTUs using Bray-Curtis dissimilarity. The impact of the various clinical diagnosis, gender, CLO test results and age of patients on community structures was determined using permutation-al ANOVA (PERMANOVA) with 1,000 permutations. Pairwise β-diversity indices were calculated using the vegdist function and then the adonis function from vegan [16]. Similarities between community structures were further visualised using non-metric multidimensional scaling (nMDS) with Bray-Curtis distance. The dispersion of the groups was determined using the betadisp function.

### Statistical analysis of patient metadata

Statistical analysis was conducted using R V4.0.2. The Shapiro-Wilk test was used to test the normality of the data. The student t-test was performed to compare differences in age between the groups. Fisher’s exact test was then used to determine differences in categorical variables (Gender and CLO test) of both groups. A *p*<0.05 was considered significant.

## RESULTS

### Demographics and clinicopathological characteristics of patients

Table 1 describes the characteristics of the patients that participated in the study. No significant difference was observed between the two groups across the various variables. Most jaundiced patients presented with underlying choledocholithiasis (11/19), followed by head of pancreas mass (4/19), which together constituted 78.9% of the jaundiced group. *Helicobacter pylori* was prevalent in 36.8% and 10% of the jaundiced and control groups, respectively.

**Table 1:**
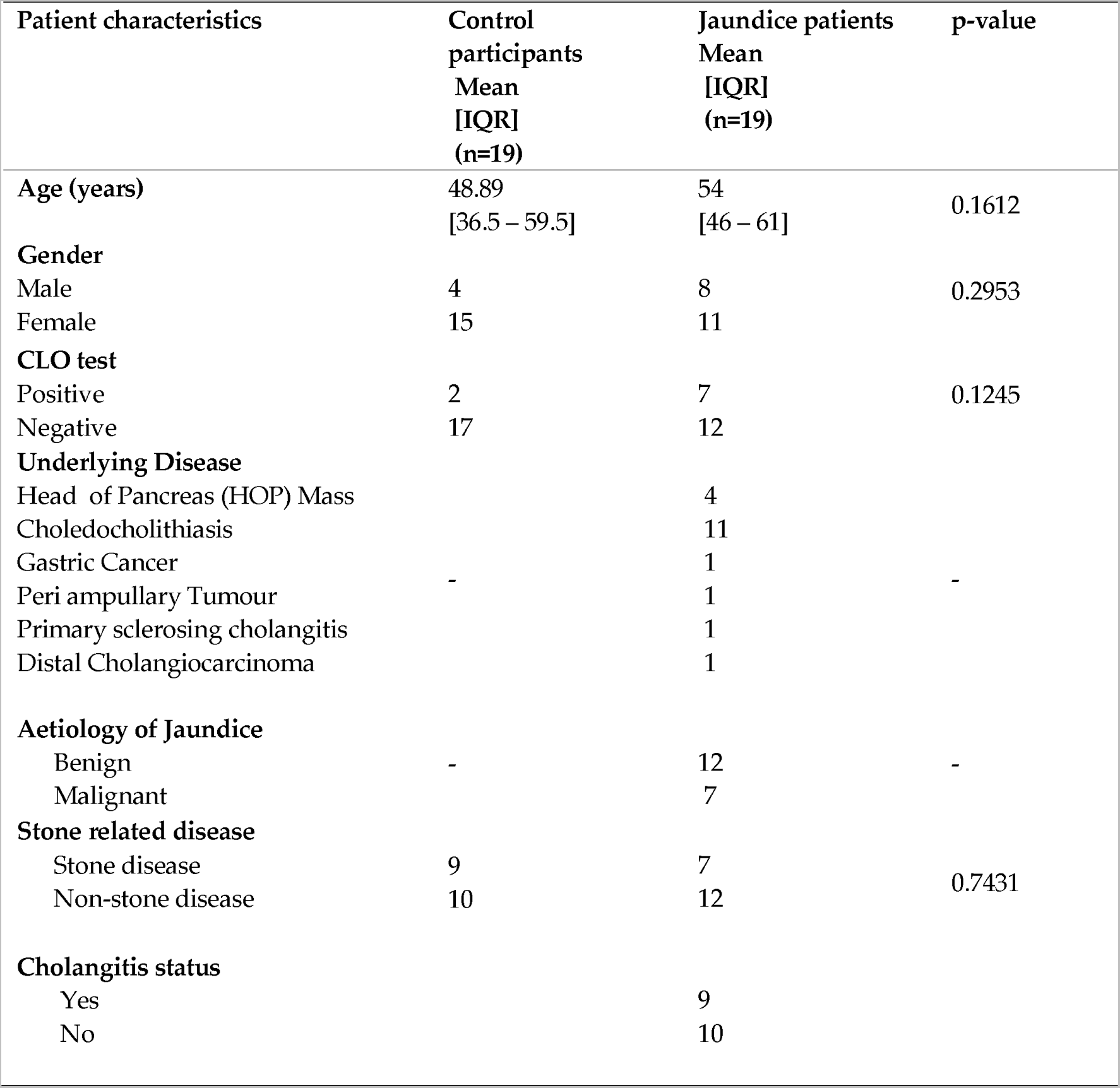
Characteristics of the Obstructive Jaundice and Control patient cohorts, and underlying etiology of jaundice

| Patient characteristics | Control participants<br>Mean<br>[IQR]<br>(n=19) | Jaundice patients<br>Mean<br>[IQR]<br>(n=19) | p-value |
| --- | --- | --- | --- |
| Age (years) | 48.89<br>[36.5 – 59.5] | 54<br>[46 – 61] | 0.1612 |
| **Gender** |  |  |  |
| Male | 4 | 8 | 0.2953 |
| Female | 15 | 11 |  |
| **CLO test** |  |  |  |
| Positive | 2 | 7 | 0.1245 |
| Negative | 17 | 12 |  |
| **Underlying Disease** |  |  |  |
| Head of Pancreas (HOP) Mass |  | 4 |  |
| Choledocholithiasis |  | 11 |  |
| Gastric Cancer |  | 1 |  |
| Peri ampullary Tumour | - | 1 | - |
| Primary sclerosing cholangitis |  | 1 |  |
| Distal Cholangiocarcinoma |  | 1 |  |
| **Aetiology of Jaundice** |  |  |  |
| Benign | - | 12 | - |
| Malignant |  | 7 |  |
| **Stone related disease** |  |  |  |
| Stone disease | 9 | 7 | 0.7431 |
| Non-stone disease | 10 | 12 |  |
| **Cholangitis status** |  |  |  |
| Yes |  | 9 |  |
| No |  | 10 |  |

### Taxonomic distribution of bacteria in the duodenum of jaundiced and non-jaundiced patients

The amplicon datasets derived from the control and jaundice groups were first rarefied to a total of 37,953 reads/patient, reflecting the lowest count present in sample S009 from a control patient. Rarefaction curves generated for each patient sample gradually reached a plateau (Figure S1), which indicates that data integrity was maintained during sample count standardisation.

Following rarefaction, a total of 1,442,214 sequences were used for the 38 samples. The bacterial communities from the 38 patient samples were classified into 14 phyla, 34 classes, and 356 genera. Bacterial phyla across both groups were predominated by Pseudomonadota (51%; mostly in the classes Gammaproteobacteria 26.66%, Betaproteobacteria 13.79% and Alphaproteobacteria 5.02%), Bacillota (17%; mainly from the classes Bacilli, 10.38%, and Negativicutes, 4.40%), Actinomycetota (16%; mostly from the class Actinomycetia, 16.13%), and Bacteroidota (13%; mostly from the class Bacteroidia 9.91%). When comparing between jaundice and control groups, variations in the relative abundance of bacterial phyla could be observed (Figure 1). The control group had a greater abundance of Pseudomonadota (54%) and Actinomycetota (17%) compared to the jaundice group (48% and 16%, respectively). In contrast, the jaundice group had a greater abundance of Bacillota (17%) and Bacteroidota (15%) compared to the control group (16% and 11%, respectively).

**Figure 1:**
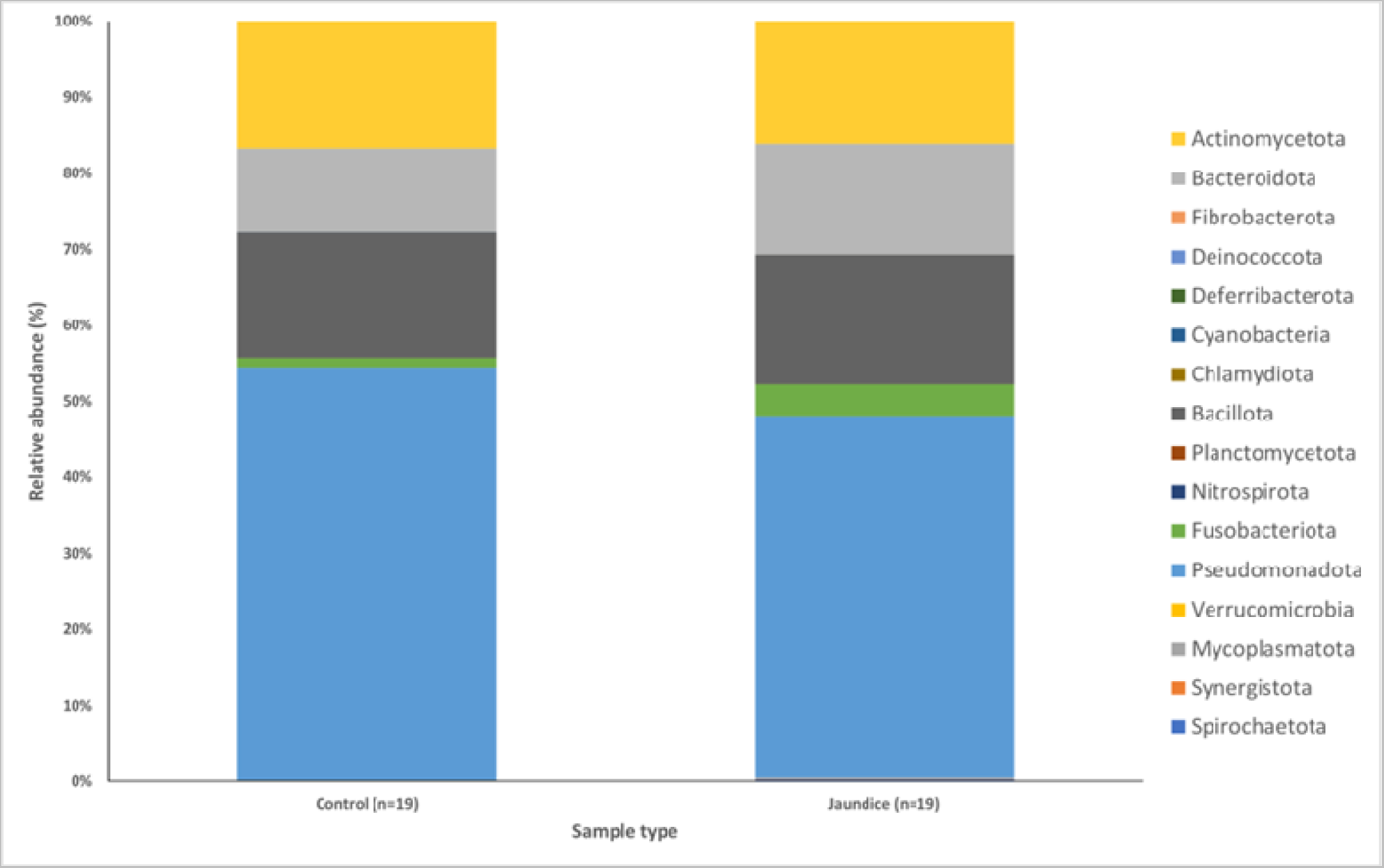
Relative abundances of the bacterial phyla found within the duodenal mucosa, obtained by biopsy during endoscopic procedures, in the control and jaundice patients. Bar chart represents the most abundant bacterial sequences classified at the phylum level in the control and jaundice cohorts.

**Figure 2:**
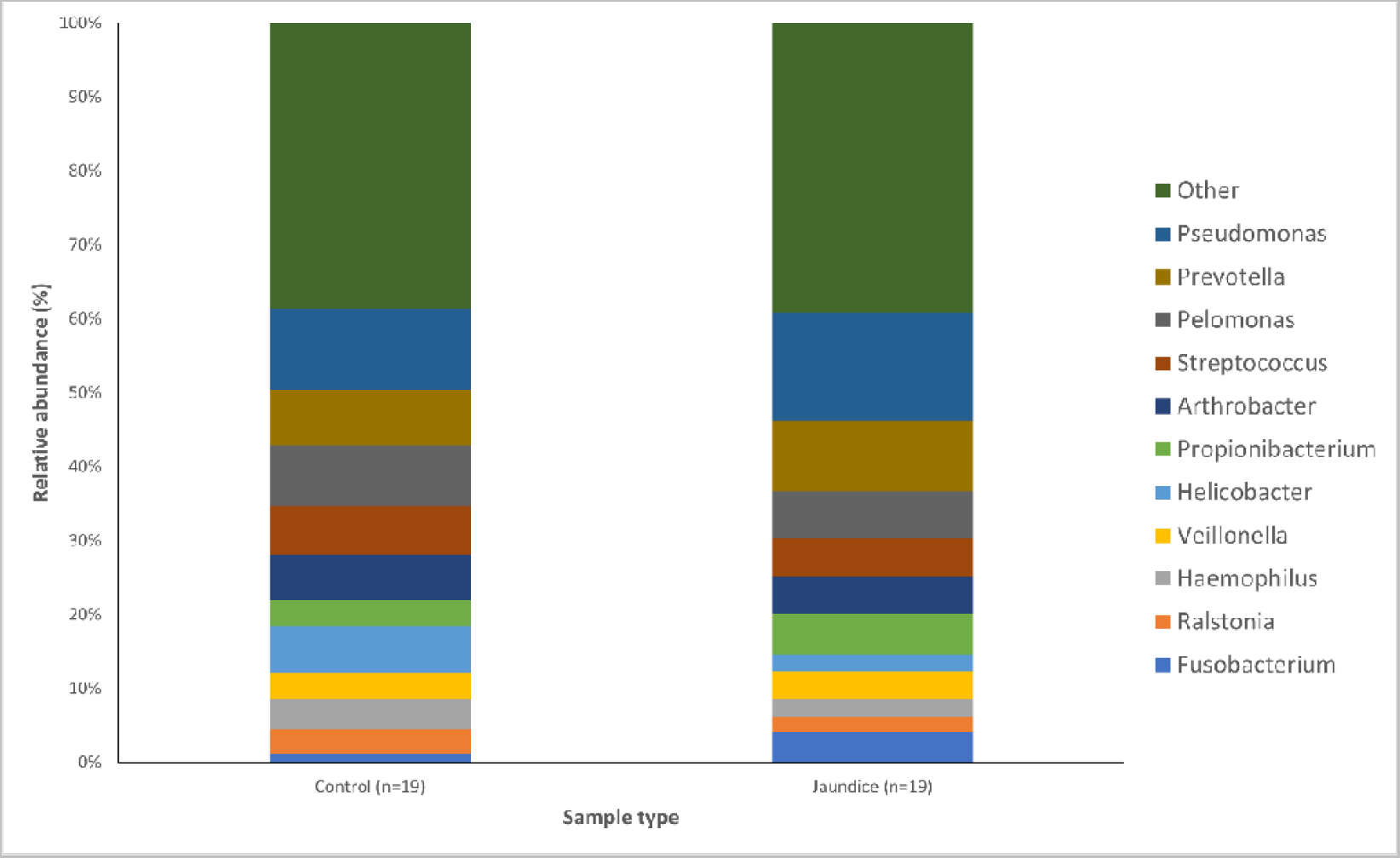
Relative abundances of the bacterial genera found within the duodenal mucosa, obtained by biopsy during endoscopic procedures, across the control and jaundice patients. Bar chart represents the most abundant bacterial sequences (> 5%) classified at the genus level in the control and jaundice cohorts. “Other” represents all other genera with relative abundance < 5%.

Variations in bacterial genera were also noted between the two groups. The control group had a greater abundance of *Pelomonas* (4.14%) and Helicobacter (3.11%) compared to the jaundice group (3.21% and 1.18%), respectively. The jaundice group in contrast had a greater abundance of *Pseudomonas* (7.29%) and *Prevotella* (4.77%) compared to the control group (5.51% and 3.71%, respectively).

### Differences in the abundance of the major bacterial phyla in the jaundiced and control group

The distribution of the data was evaluated using the Shapiro-Wilk test observed to be normally distributed. Multivariant testing using Dirichlet-Multinomial distribution was performed to test for differences in the mean distribution of the seven most abundant bacterial phyla. While there was an observed difference in the absolute abundances of these bacteria (Figure 1) between the control and jaundiced groups, there was no significant difference in the mean distribution of these taxa between jaundice and control groups (Chi-square = 14.87 and *p*= 0.09). In general, Pseudomonadota, Bacillota, Bacteroidota and Actinomycetota constituted the predominated phyla across all the specimens irrespective of the underlying condition. Nevertheless, multivariate analysis showed a statistically significant difference between patients with cholelithiasis in the control group and those with a head of pancreas mass in the jaundiced group (Chi-square = 20.31 and *p* <0.01) (Figure 3).

**Figure 3:**
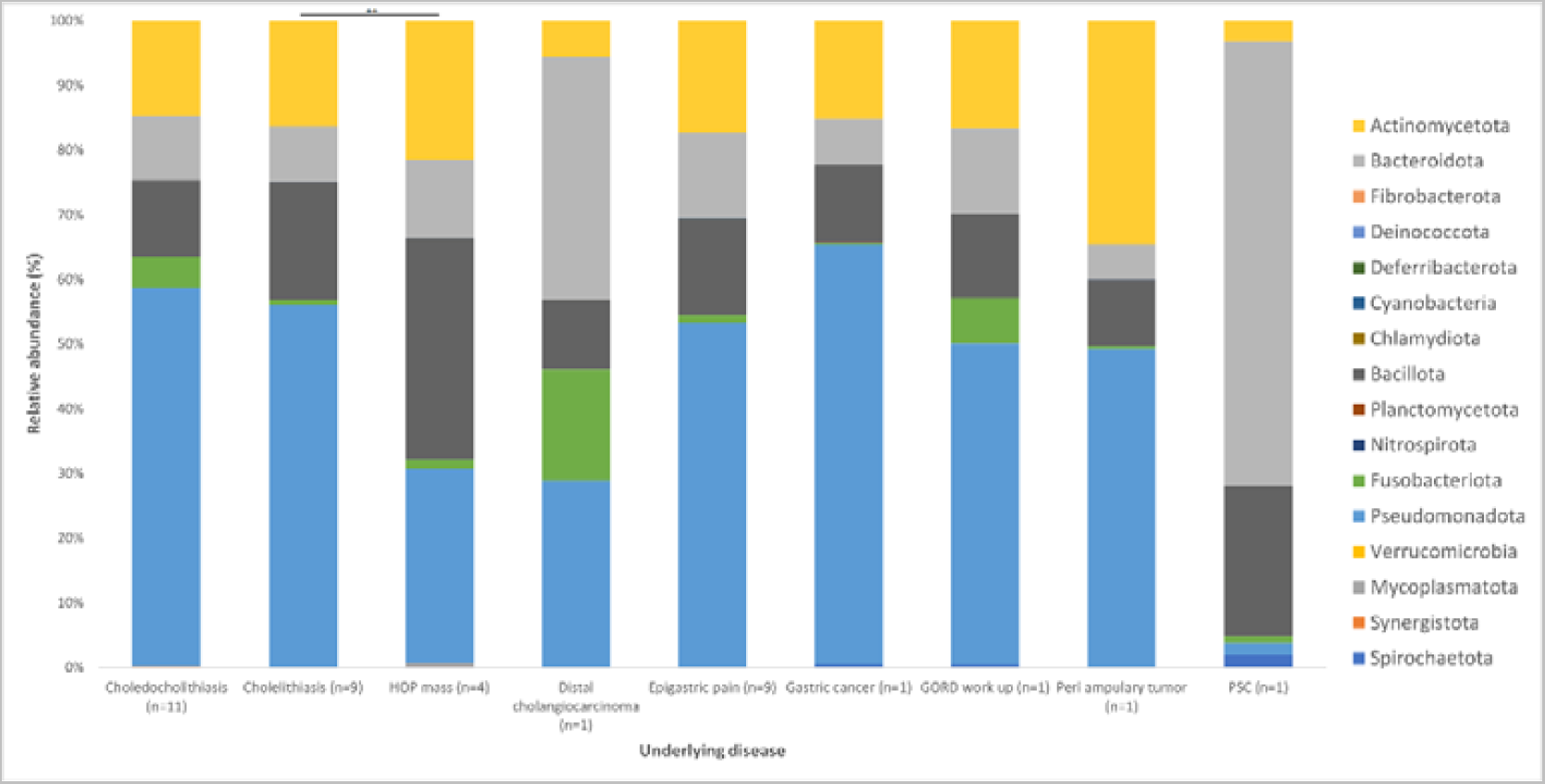
Relative abundances of bacterial phyla found within the duodenal mucosa across variable demographic data points collected between the control and jaundiced groups. The bar chart represents the most abundant bacterial sequences classified at the phylum level in the control and jaundice cohorts classified according to the underlying disease presented by the individual patients. A significant difference in the mean distribution of the seven most abundant phyla was noted between patients with cholelithiasis and patients with head of pancreas (HOP) mass (p< 0.01). Differences in mean distribution was tested with a multivariant test using Dirichlet-Multinomial distribution.

Patients with cholelithiasis demonstrated a greater abundance of *Pseudomonas* (21.14%) with samples S004, S005, S007 and S015 accounting for 18.67% of this total and *Helicobacter* (4.42%) with sample S010 accounting for 4.01% of this total compared to patients with HOP mass (4.66% and 0.03%, respectively) with only patient samples S027 and S033 showing the presence of Helicobacter.

HOP mass patients in contrast demonstrated a greater abundance of *Veillonella* (12.46%) and *Propionibacterium* (7.24%) compared to cholelithiasis patients (5.22% and 3.43%, respectively). However, while *Veillonella* was found to be prevalent in eight of the cholelithiasis patients, only two HOP mass patients presented with taxa in this genus, with sample S020 further accounting for 11.13% of the total abundance of *Veillonella* in HOP mass patients. While *Staphylococcus* was present across all samples of the cholelithiasis (0.78%) and HOP mass (3.21%) cohort, patient samples S020 and S031 with HOP mass accounted for 2.90% towards the total abundance of this species within HOP mass patients. Furthermore, *Bergeyella* was found to be present in only two out of the four HOP mass patients compared to no patients with cholelithiasis, with patient sample S033 accounting for 3.59% of the total abundance of this species. This variation in the relative abundance of bacterial species in the two cohorts may be linked to the overrepresentation of the HOP mass cohort with four samples only compared to the cholelithiasis cohort which had a total of nine samples.

**Figure 4:**
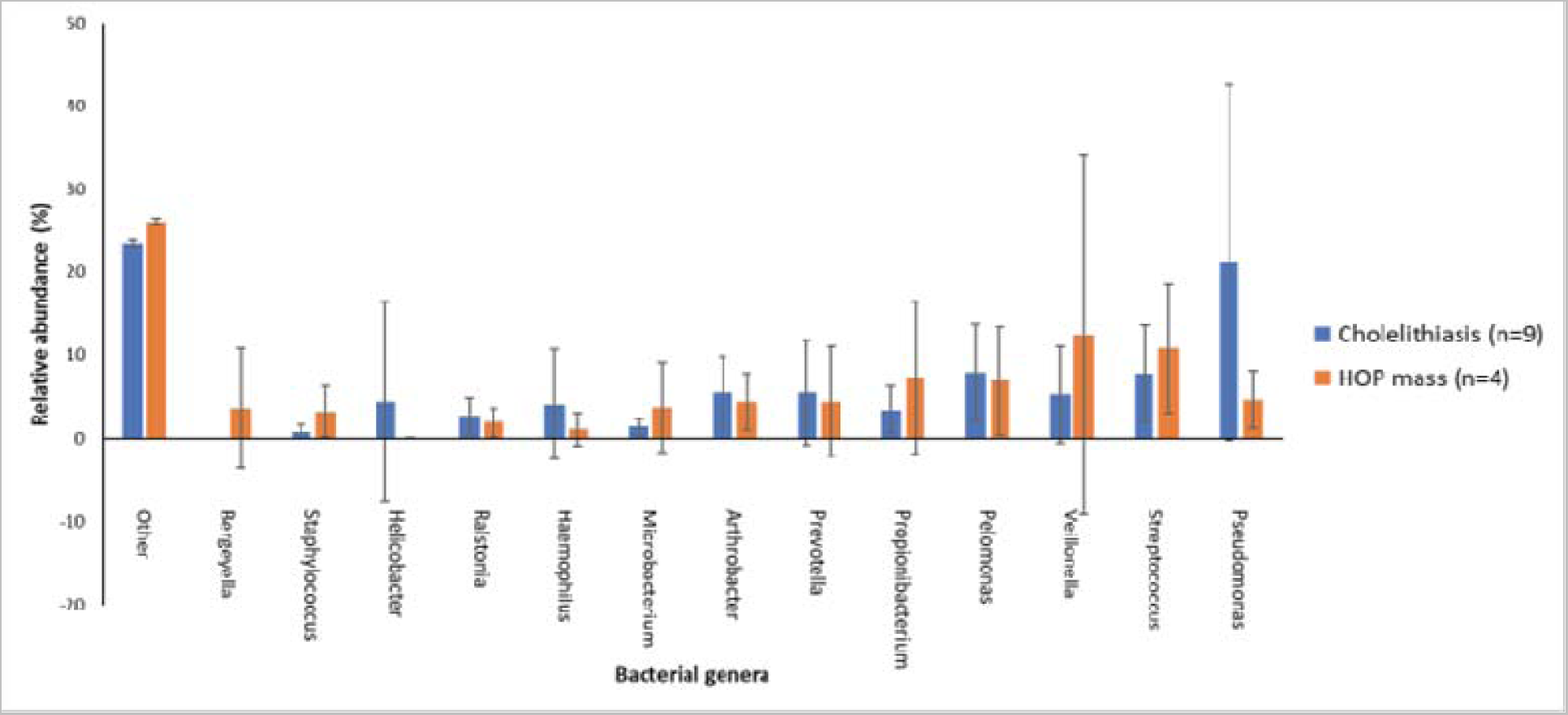
The distribution of the main bacterial genera between cholelithiasis and HOP mass patient duodenal samples. A bar chart represents the most abundant bacterial sequences (> 3.5%) classified at the genus level in patients with cholelithiasis or HOP mass as an underlying disease. “Other” represents all other genera present within these patients with relative abundance < 3.5%. Patients with cholelithiasis demonstrated a greater abundance of *Pseudomonas* and *Helicobacter* compared to patients with HOP mass. In contrast, HOP mass patients demonstrated a greater abundance of *Veillonella* (12.46%) and *Propionibacterium* (7.24%) compared to cholelithiasis patients (5.22% and 3.43%, respectively).

### Evaluation of the mean distribution of bacteria when comparing patients with cholangitis vs no cholangitis

Patients were determined to have cholangitis based on the widely accepted criteria of the Tokyo guidelines of 2018 (TG18). In the jaundiced cohort of patients, 9 patients had cholangitis while 10 patients did not. The mean distributions of the organisms at both genus and phylum levels were compared between jaundiced patients with cholangitis, those without cholangitis and the control group of patients. A statistically significant difference existed when comparing the jaundiced group with cholangitis versus those without cholangitis (*p*=0.0017) as well as when comparing the cholangitis group to the control group (*p*= 0.0026).

**Figure 5:**
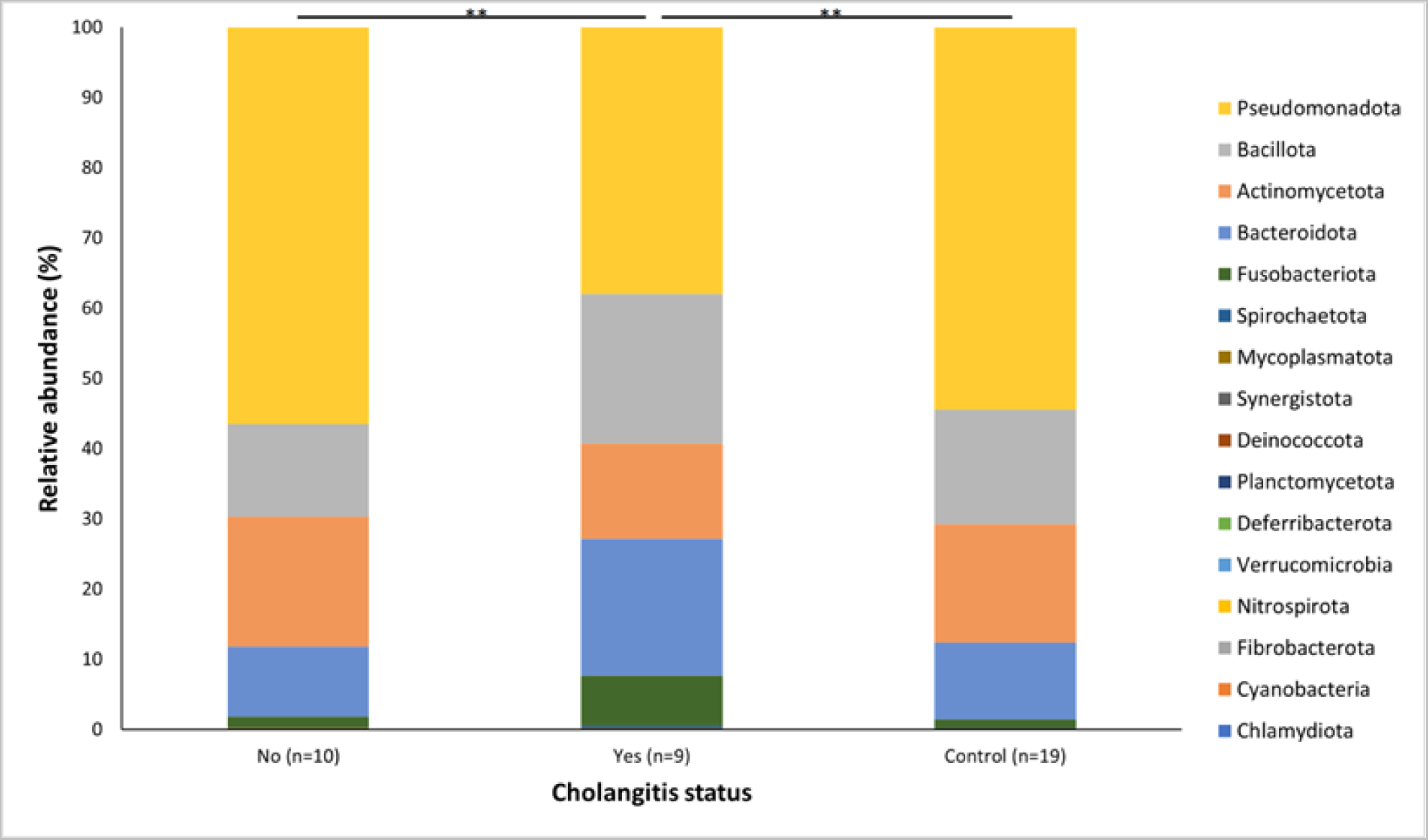
Relative abundances of the bacterial genera found within the duodenal mucosa, comparing the control group and jaundice group. The jaundiced group was further subdivided into those with cholangitis (Yes) and without cholangitis (No). A Bar chart represents the most abundant bacterial sequences (> 4.5%) classified at the genus level in the control and jaundice cohorts. “Other” represents all other genera with relative abundance < 4.5%. ** denotes p< 0.01

At the phylum level, the group of patients with cholangitis had an increased abundance of Bacteroidota (19.49%) and Fusobacteriota (7.01%) compared to the control group (11.00% and 1.27%) and the jaundice group without cholangitis (10.01% and 1.48%), respectively. By contrast, both the control group and jaundiced group without cholangitis had a greater prevalence of Pseudomonadota (54.34% and 56.42%) as well as Actinomycetota (16.79% and 18.53%), compared to the patients with cholangitis (37.97% for Pseudomonadota and 13.57% for Actinomycetota, respectively).

**Figure 6:**
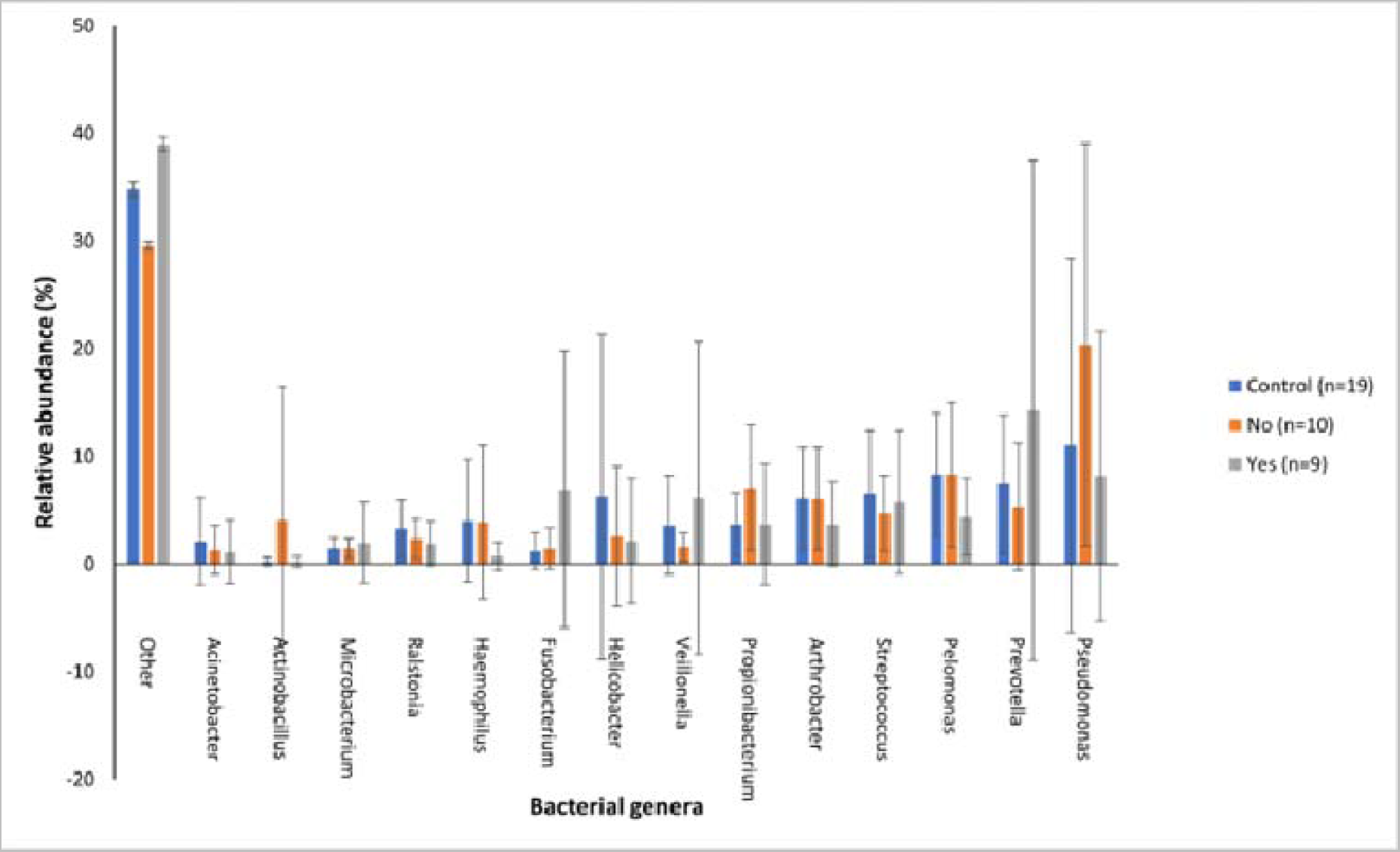
The distribution of the main bacterial genera between the control group and the jaundiced group, which was divided into two subgroups, those with cholangitis (Yes) and those without cholangitis(No). Bar chart represents the most abundant bacterial sequences (> 4.5%) classified at the genus level in patients with cholelithiasis and HOP mass as an underlying disease. “Other” represents all other genera present within these patients with relative abundance < 4.5%.

The jaundiced group with cholangitis had an increased abundance of three Gram-negative genera, *Prevotella* (14.27%), *Fusobacterium* (6.92%) and *Veillonella* (6.11%) when compared to the control group (7.42%, 1.19% and 3.62%) and jaundiced group without cholangitis (5.30%, 1.43% and 1.57%), respectively. While the cholangitis group had a decreased abundance of the Gram-negative genus *Pseudomonas* (8.16%) and the Gram-positive genus *Propionibacterium* (3.69%) when compared to the control group (11.02% and 3.69%) and the jaundiced group without cholangitis (20.37% and 7.10%), respectively.

### Evaluation of bacterial **α**-diversities among jaundiced and non-jaundiced patients

To evaluate the diversity of bacterial species in the duodenum of both jaundiced and control patients based on various variables, the observed OTU richness and Shannon-Weaver diversity indices were determined. A slightly higher richness was observed for the nineteen control samples compared to the nineteen jaundice samples for observed OTU richness (Figure 7C). Similarly, when comparing the alpha diversity based on CLO test outcomes, a slightly higher OTU richness was noted for the CLO-negative patients compared to the CLO-positive patients. This could have resulted due to a greater classification of samples with a CLO-negative (twenty-nine samples) outcomes compared to CLO-positive (nine samples) outcomes (Figure 7A). Comparison of the bacterial α-diversities of duodenal samples for patients with the stone disease (sixteen patients) and those with non-stone disease (twenty-two patients) showed greater between-sample variability in diversity for those patients with the non-stone-related disease (Figure 7B). The relative abundance of Pseudomonadota is increased in patients with stone-related disease compared to those with non-stone-related disease (54.6% and 46.5%, respectively) (Table S4). The phylum Actinomycetota is comparatively overrepresented in patients with non-stone disease as compared to those with stone-related disease (18.4% and 14.8%, respectively). When comparing the α-diversity of duodenal bacterial communities in jaundiced patients with underlying benign diseases (twelve patients) as compared to patients with underlying malignant diseases (seven patients), greater variability is observed among the benign disease cohort (Figure 7D). Patients with malignant causes of jaundice had an increased prevalence of Actinomycetota (20.2%) and Bacillota (24.2%) when compared to patients with benign causes for jaundice (13.8% and 12.8%, respectively) as well as compared to patients in the control group (16.7% and 16.4%, respectively) (Table S5).

### Assessment of the influence of multifactorial metadata on beta diversity

Bacterial communities across patients’ samples in the control cohort grouped more tightly together compared to the jaundice cohort, and were fully covered by the jaundiced cohort (Figure 8A). Patients samples from the jaundice cohort exhibited greater variability in bacterial communities with samples S029 and S036 lying outside of the main clusters of the two cohorts while only sample S018 from the control cohort seemed to lie outside of the main clusters from the control cohort. Similar to the distribution of bacterial community structures in jaundice and control cohorts, patient samples with non-stone disease seemed to cluster more closely together compared to those with stone disease (Figure 8B). These non-stone disease samples were further fully overlapped by those with stone disease. While patient samples with stone disease exhibited greater variability in bacterial communities with samples S029 and S023 lying outside of the main cluster, patient samples S018 and S020 from the non-stone dis-ease samples also exhibited variation from the main cluster and lay outside of the stone disease and non-stone disease clusters. When considering the bacterial community structures across 1) CLO positive and CLO negative testing, 2) Control, benign and malignant samples and 3) across the underlying conditions of both the control and jaundiced patients, all cohorts clustered closely together (Figure S2).

**Figure 7:**
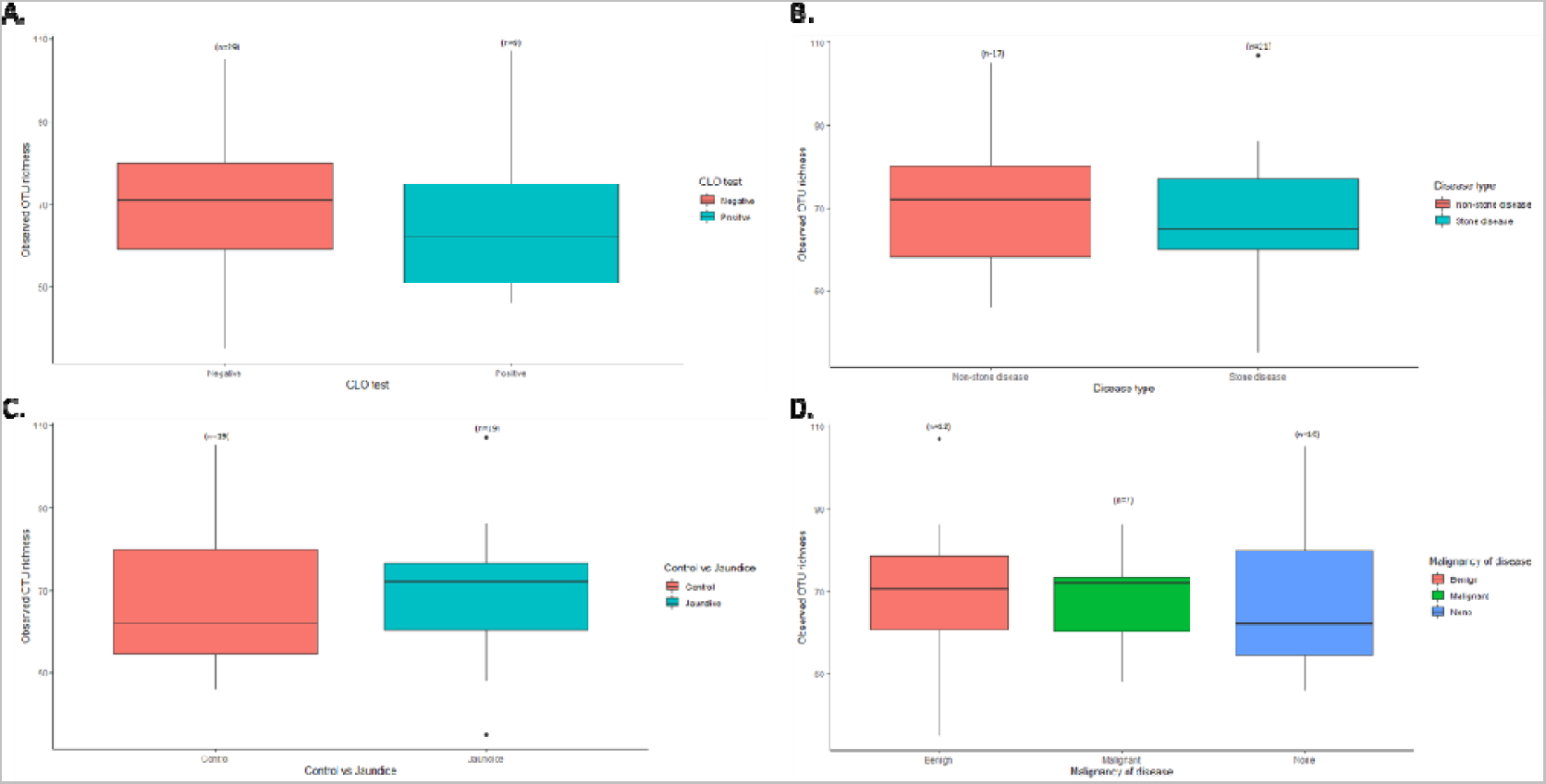
Variation in α-diversity indices for bacterial communities of the duodenal mucosa in the different cohorts. Boxplots show the observed OTU richness indices for the different clinical characteristics from both groups of patients for the bacterial taxa present in A) CLO-positive vs CLO-negative groups, B) stone-related disease vs non-stone-related disease groups C) control vs jaundice groups and D) benign vs malignant groups compared to the control. Differences between the various cohorts were evaluated using ANOVA and Tukey HSD which indicated no significant difference between the different cohorts individually and as interactions.

**Figure 8:**
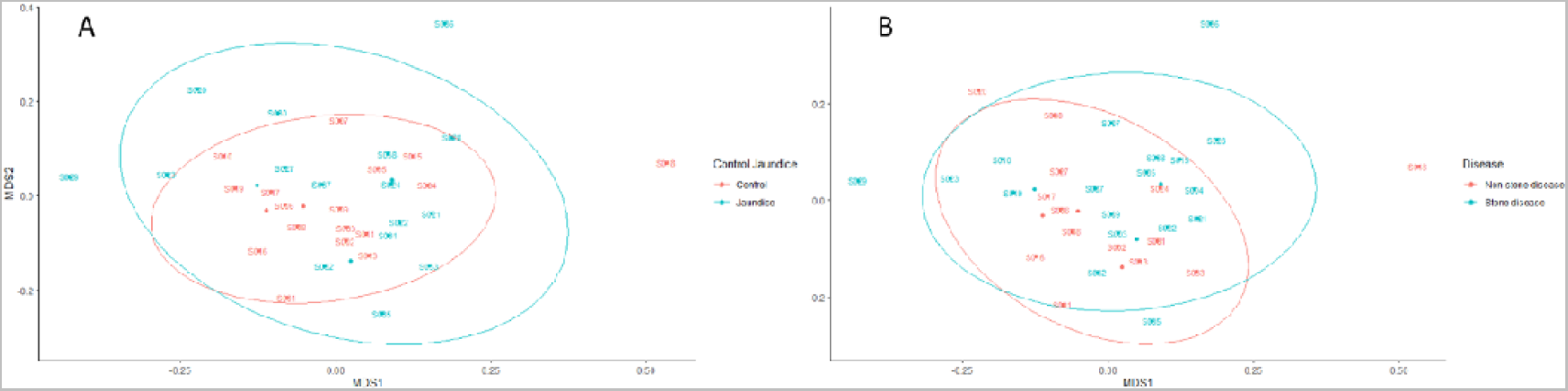
Bacterial community structures contained within the duodenal mucosa, obtained by endoscopic biopsies. nMDS plots represent the bacterial community structures amongst A) control vs jaundice and B) stone vs non-stone disease patients. The dispersion of the points is representative of the dissimilarities in community structures. The bacterial stress solution was reached at about 0.20. Differences in the variation of community structures between the control vs jaundice and stone vs non-stone disease were tested using PERMANOVA based on Bray-Curtis dissimilarity. This indicated no significant difference in the variations of bacterial community structures amongst the different cohorts individually and as interactions.

Variations in bacterial community structures for the control and jaundice cohorts were further analysed using beta dispersion. This revealed that there was no difference in the composition of bacterial communities between the two groups of patients (*p*= 0.60). This finding is demonstrated by the overlap between the control and jaundice cohorts and those with stone disease and non-stone disease (Figures 8A and 8B). However, when examining the beta diversity of the duodenal microbiome with the CLO status factored in, there was a significant difference in the bacterial OTUs between samples of patients with stone disease vs non-stone disease patients (*p* = 0.04) (Table S6).

## DISCUSSION

The gut microbiota has been associated with the normal functioning of key physiological processes throughout the body. This study aimed to determine if the obstruction of bile flow, and concomitantly jaundice, is linked to a change in the microbiome within the duodenum.

Across both jaundiced and control patients, the predominant bacterial phylum of the duodenum in our patient cohort was the Pseudomonadota (Figure 1). This contrasts with a similar study which demonstrated the dominant phylum to be Bacillota with a prevalence of 59%, with Pseudomonadota only accounting for 15% of the bacterial phyla [17]. It should be noted that the previous study looked at duodenal fluid, while mucosal biopsies were sampled in this study. Furthermore, the cohort of the previous study included children of median age of 15 with irritable bowel disease [17]. Of note in the current study, Pseudomonadota were more abundant in the control group compared to jaundiced patients. However, it was unclear what factors contribute to the abundance of this bacterial phylum. At the genus level, the control group had a larger abundance of *Pelomonas* and *Helicobacter* compared to the jaundiced group(Figure 2), while the jaundice group had an increased abundance of *Pseudomonas* and *Prevotella* compared to the control group. The abundance of *Helicobacter* observed by 16SRNA sequencing was in contrast to the rate of CLO test positivity, in which most of the control patients were negative. This contrast might be due to the sensitivity of the CLO test which varies between 60 -90% [18,19]. The increased prevalence of *Pseudomonas* in the jaundice cohort corroborates a recent finding that showed its abundance in bile aspirates of jaundiced patients [20]. Additionally, we found an increased prevalence of Actinomycetota and Bacillota in jaundiced patients due to malignancy compared to those with benign causes. Importantly, the occurrence of Bacillota has been observed in cancer patients with some of these linked to the use of catheters [21].

A multivariant test using Dirichlet-Multinomial distribution showed that patients with cholelithiasis had a significant difference in microbiota to those with a head of pancreas mass. Specifically, at the genus level, patients with cholelithiasis demonstrated a greater abundance of *Pseudomonas* and *Helicobacter* compared to patients with HOP mass. This increased prevalence of *Pseudomonas* in patients with cholelithiasis is consistent with a recent study that demonstrated that the presence of *Pseudomonas* contributes to the formation of gallstones [22]. *Helicobacter* has also been implicated in the development of cholelithiasis [23,24] As such, this study further validates the available evidence that *Pseudomonas* and *Helicobacter* are associated with the formation of cholelithiasis. Patients with HOP mass demonstrated a greater abundance of *Veillonella* and *Propionibacterium* compared to cholelithiasis patients. The prevalence of *Veillonella*, an organism generally associated with oral microbiomes, was also observed in a study of patients with pancreatic cancer [25]. *Propionibacterium* has been implicated in infections including those resulting in post-operative wounds from surgeries [26]. Since the patients used in this study underwent ERCP, the relative abundance of *Propionibacterium* may hint towards its potential role in post-operative infections, which is a problem of significant concern in patients undergoing major pancreatic surgery after ERCP [26].

To the best of our knowledge, this is the first study that utilizes next-generation sequencing (NGS) to examine the difference between the duodenal microbiota in jaundiced patients with cholangitis and jaundiced patients without cholangitis. Cholangitis is a life-threatening condition that has been linked to the migration of bacteria from the gastrointestinal tract into the usually sterile biliary tract [24,27]. In this study, we demonstrated a significant difference in the distribution of bacteria in jaundiced patients with cholangitis compared to those without cholangitis. Patients with cholangitis demonstrated an increased prevalence of *Prevotella*, *Fusobacterium* and *Veillonella*. *Prevotella* was found to be the most abundant anaerobic organism isolated from patients undergoing liver resection who developed infected post-operative bilomas [28].Therefore, it is suggested that *Prevotella* may be an important role in biliary sepsis with implications for targeting it for treatment. *Fusobacterium* is an uncommon infectious organism, however, when present is associated with significant mortality rates [29,30]. It has been demonstrated to be an important causative organism of biliary sepsis (e.g. in emphysematous cholecystitis) [30]. *Fusobacterium* is seldomly identified using standard culture techniques and as such NGS offers an ideal solution for the detection of this potentially lethal organism. It has been demonstrated to be an important causative organism for biliary sepsis cases such as emphysematous cholecystitis [30]. *Veillonella* is a normal commensal organism but becomes an opportunistic pathogen when overgrown [31]. *Veillonella* tends to be over-represented in populations where there is a reduced flow of bile into the duodenum [32]. This would explain its presence in patients with obstructive jaundice and its over-representation in patients with cholangitis.

The comparison of the alpha diversities across the different underlying conditions showed multiple significant results, however, they were comparing a single patient with PSC to larger patient groups with cholelithiasis, choledocholithiasis, head of pancreas mass or patients with epigastric pain (Table S9). It is important to note that PSC has been shown to affect the mucosal barrier of the GIT, increasing its permeability, and affecting the enterocytes from carrying out base functions, such as nutrient absorption and immune modulation [31]. Future research should further investigate changes in the microbiota in a larger cohort of patients with PSC.

Bray-Curtis measure of dissimilarity of the beta diversities demonstrated a significant difference between duodenal microbiota when comparing patients with stone-related disease to those patients without and factoring in the outcomes of the CLO test (Table S6). The abundance of Pseudomonadota is increased in patients with stone-related disease compared to those with non-stone-related dis-ease. There is an increasing body of evidence that demonstrates that the gut microbiota can contribute to the development of specific types of gallstones (cholesterol, pigmented or mixed types) [23]. Furthermore, the differences between the 2 groups were only observed when considering the result of the CLO test, corroborating the potential role of *Helicobacter pylori* in the development of gallstones [23].

## CONCLUSION

The GIT microbiome is diverse and has been linked to many pathological conditions. In this pilot study, we aimed to contribute to an understudied area of microbiomes, that of the duodenum when jaundice is present. We showed some differences in the duodenal microbiota between jaundiced and non-jaundiced patients, although these were not significant. However, when considering the underlying aetiology of jaundice significant differences were identified. Of importance, there existed a significant difference between jaundiced patients with cholangitis compared to those without cholangitis. It is still unclear if the observed shifts are causal or a consequence of the different disease conditions. In the future, larger studies would add to these findings and help in elucidating the role of the key microbes in the pathophysiology of these conditions.

## Data Availability

All data produced in the present study are available upon reasonable request to the authors

## SUPPLEMENTARY TABLES AND FIGURES

### SUPPLEMENTARY FIGURES

**Figure S1:**
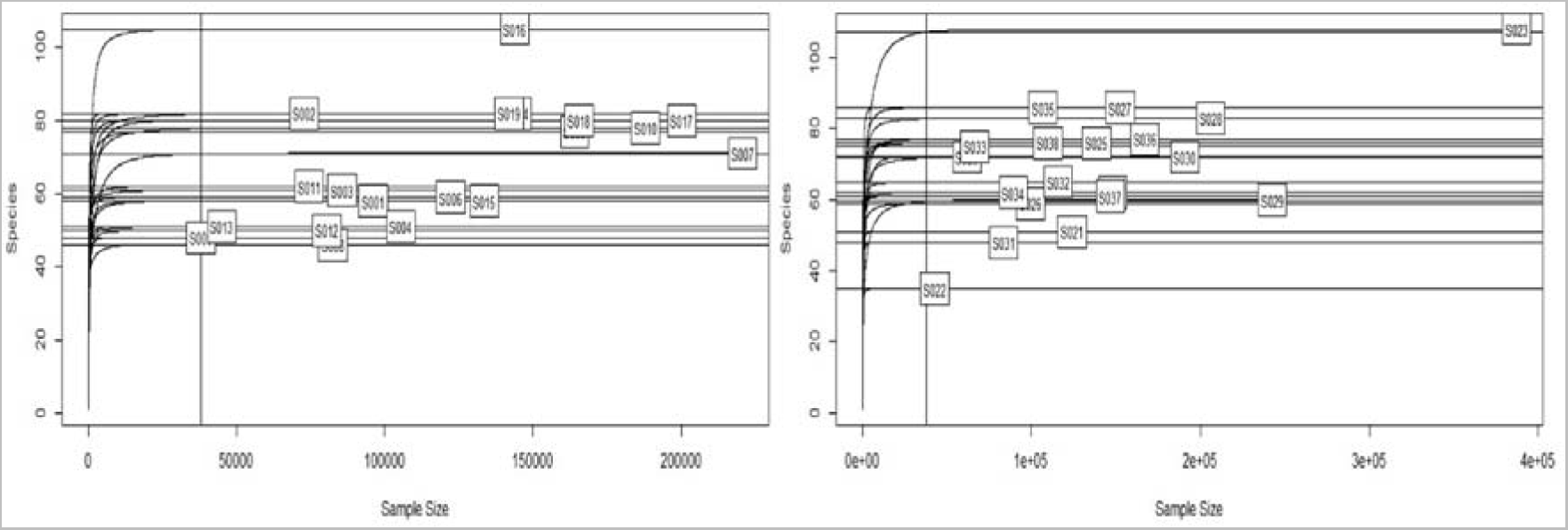
Bacterial rarefication curves, rarefied to 37,953 OTUs representing the lowest count in sample 9 of the control group.

**Figure S2:**
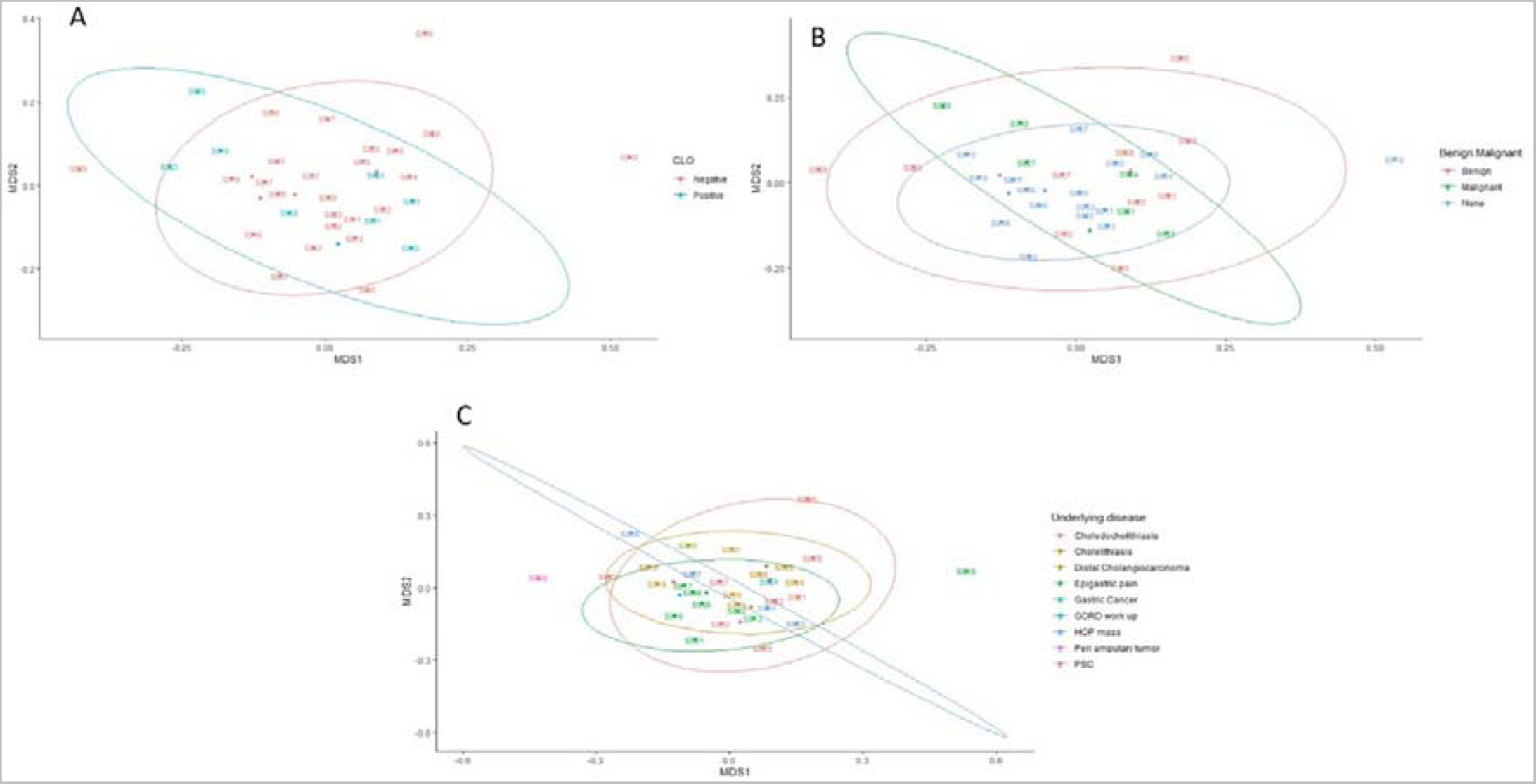
Bacterial community structures represented as nMDS plots across. A) CLO positive and CLO negative testing B) Control, benign and malignant samples and lastly C) across the underlying conditions of both the control and jaundiced patients. The distance between points is indicative of the relative dissimilarities in community structures with the ellipses indicating the dispersion regions of each site. Bacterial stress solution was reached at about 0.20 which was relatively low.

### SUPPLEMENTARY TABLES

**Table S1:**
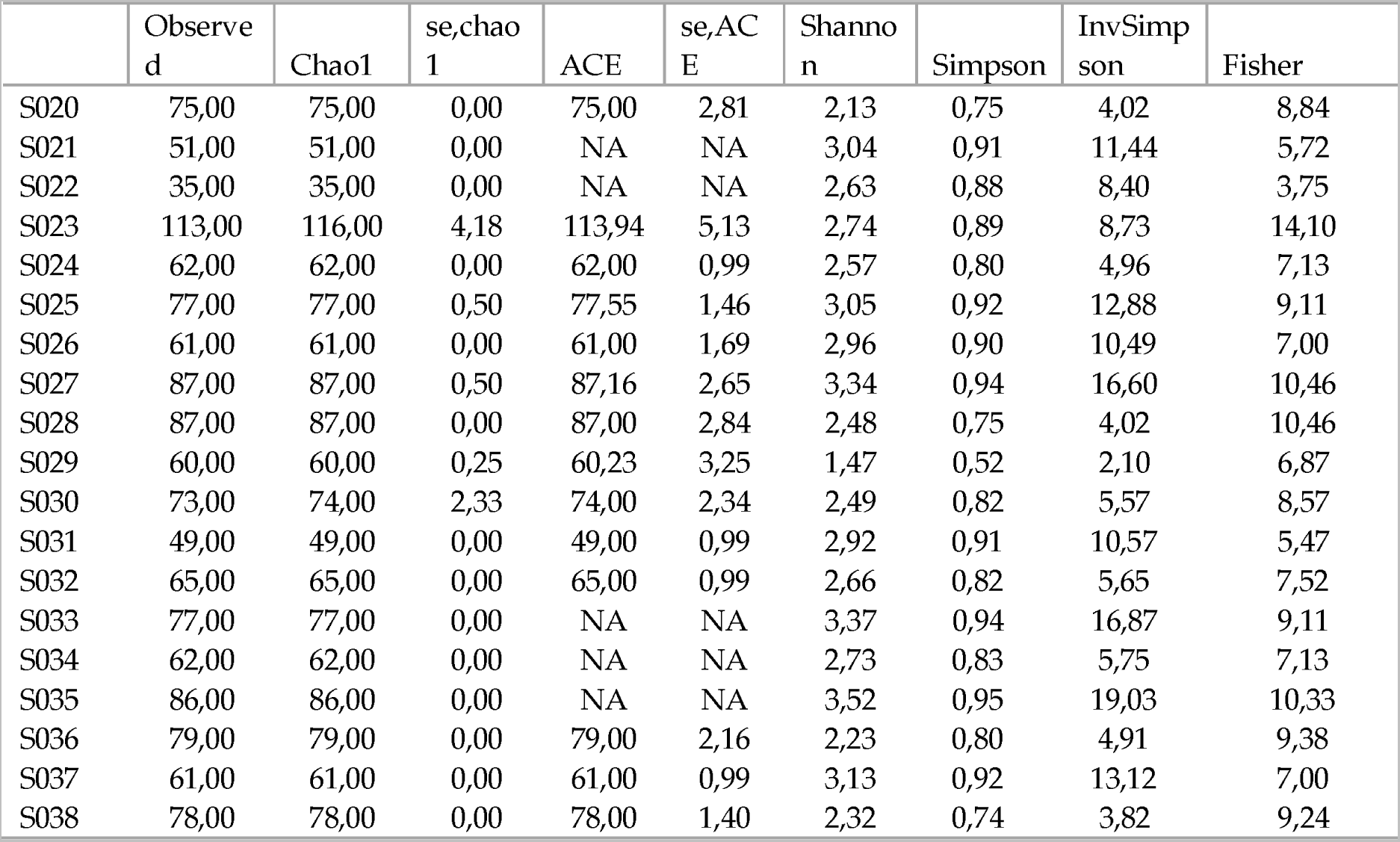
Diversity indices of individual samples in the jaundice group

**Table S2:**
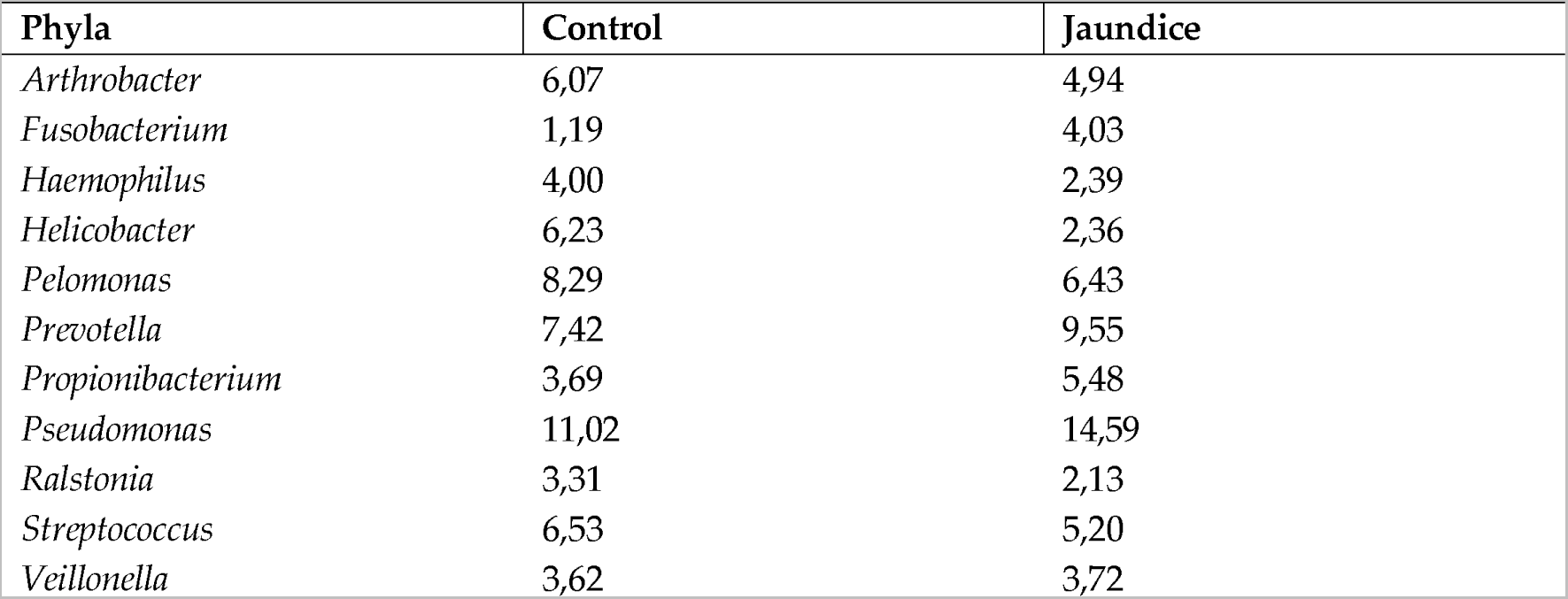
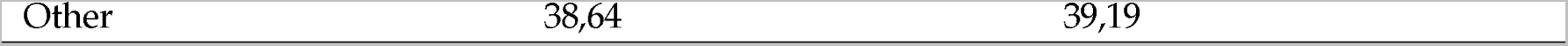
Mean distribution of phyla comparing control and jaundice cohorts

**Table S3:**
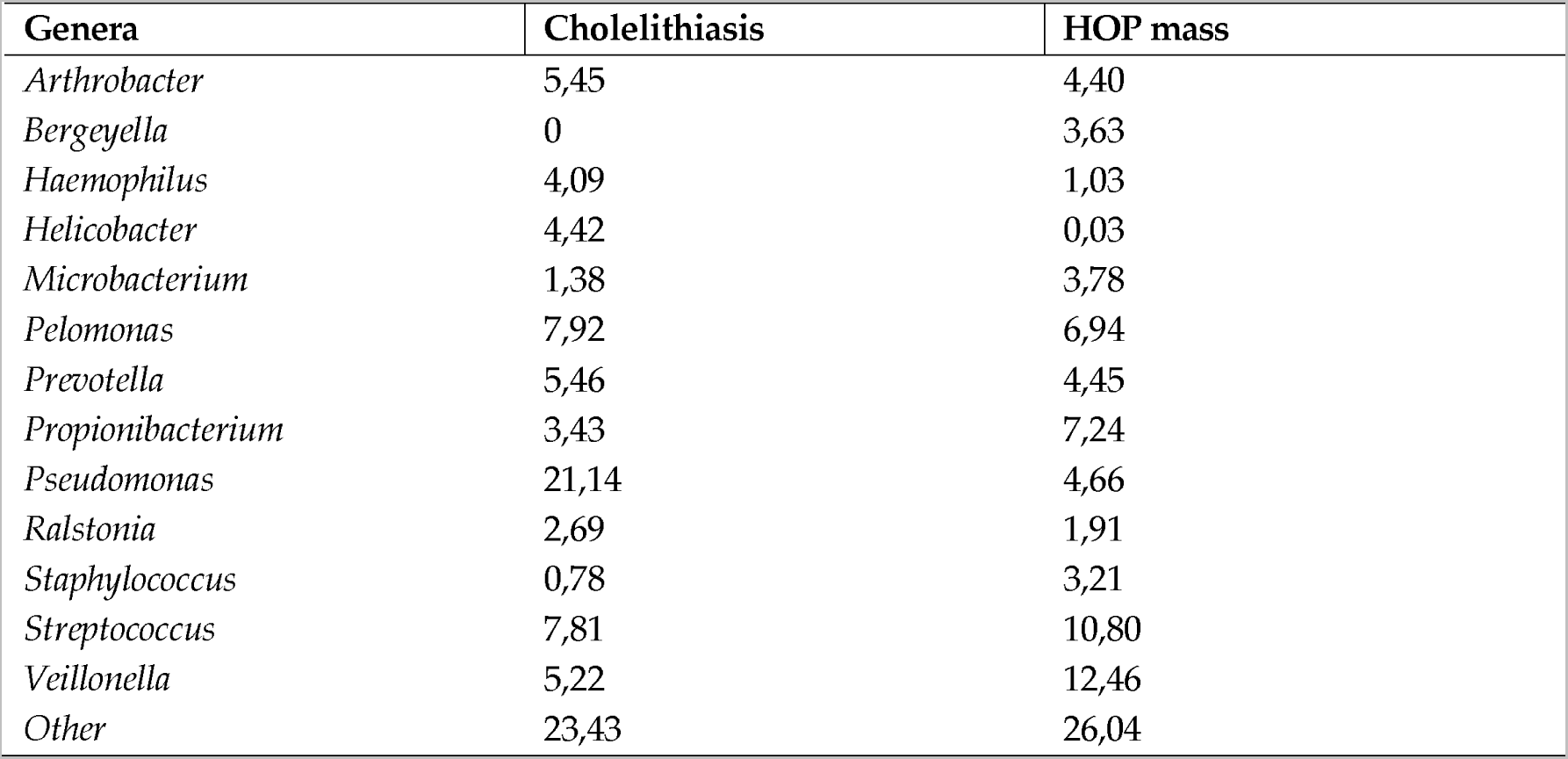
Mean distribution of genera comparing patients with cholelithiasis and HOP (head of pancreas) mass

**Table S4:**
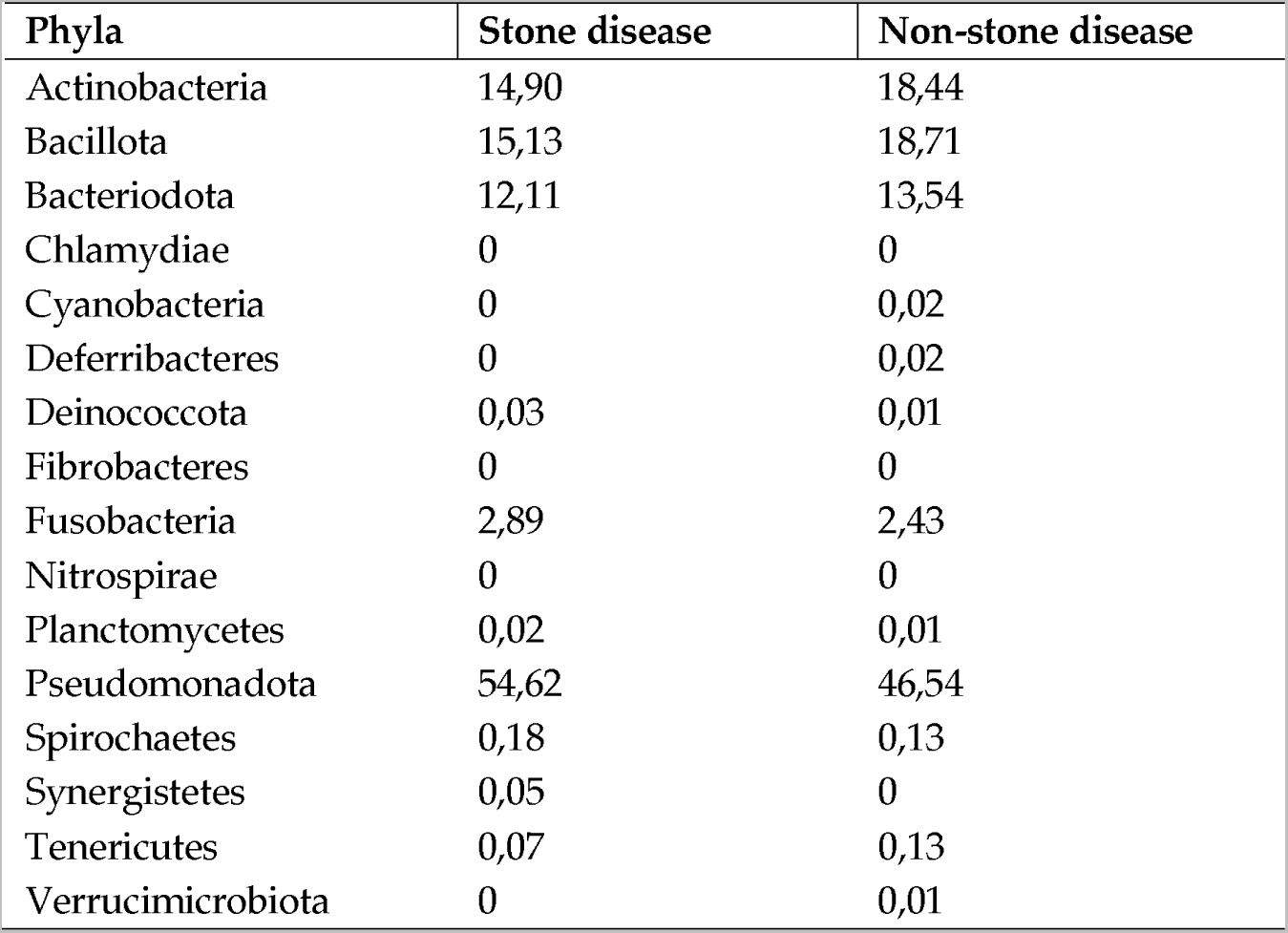
Relative abundances, expressed as a percentage, of the bacterial phyla in the duodenum of patients when grouped according to the presence or absence of stone disease.

**Table S5:**
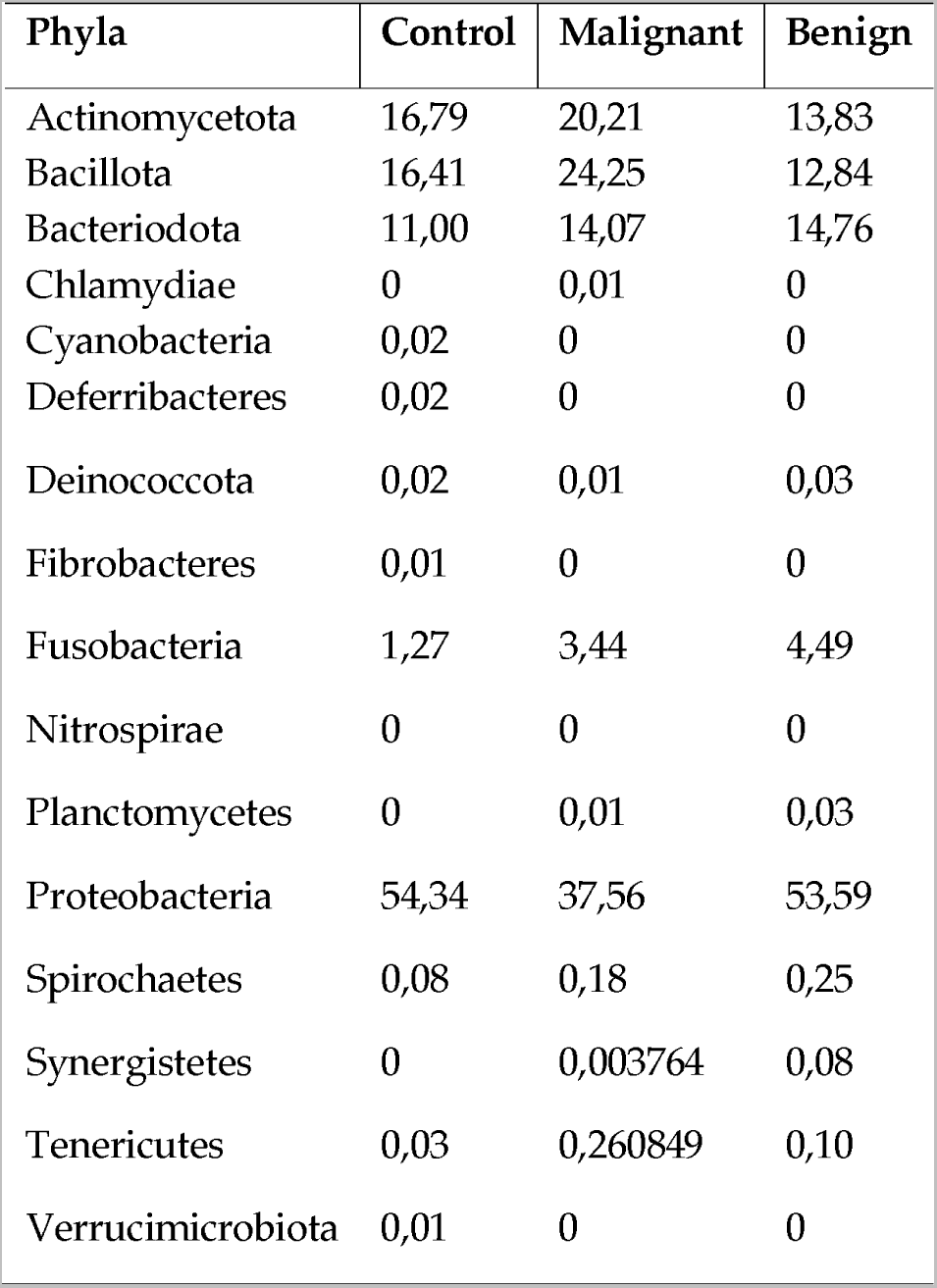
Relative abundances, expressed as a percentage, of the bacterial phyla in the duodenum of patients, comparing patients with benign and malignant conditions for jaundiced as well as control patients

**Table S6:**
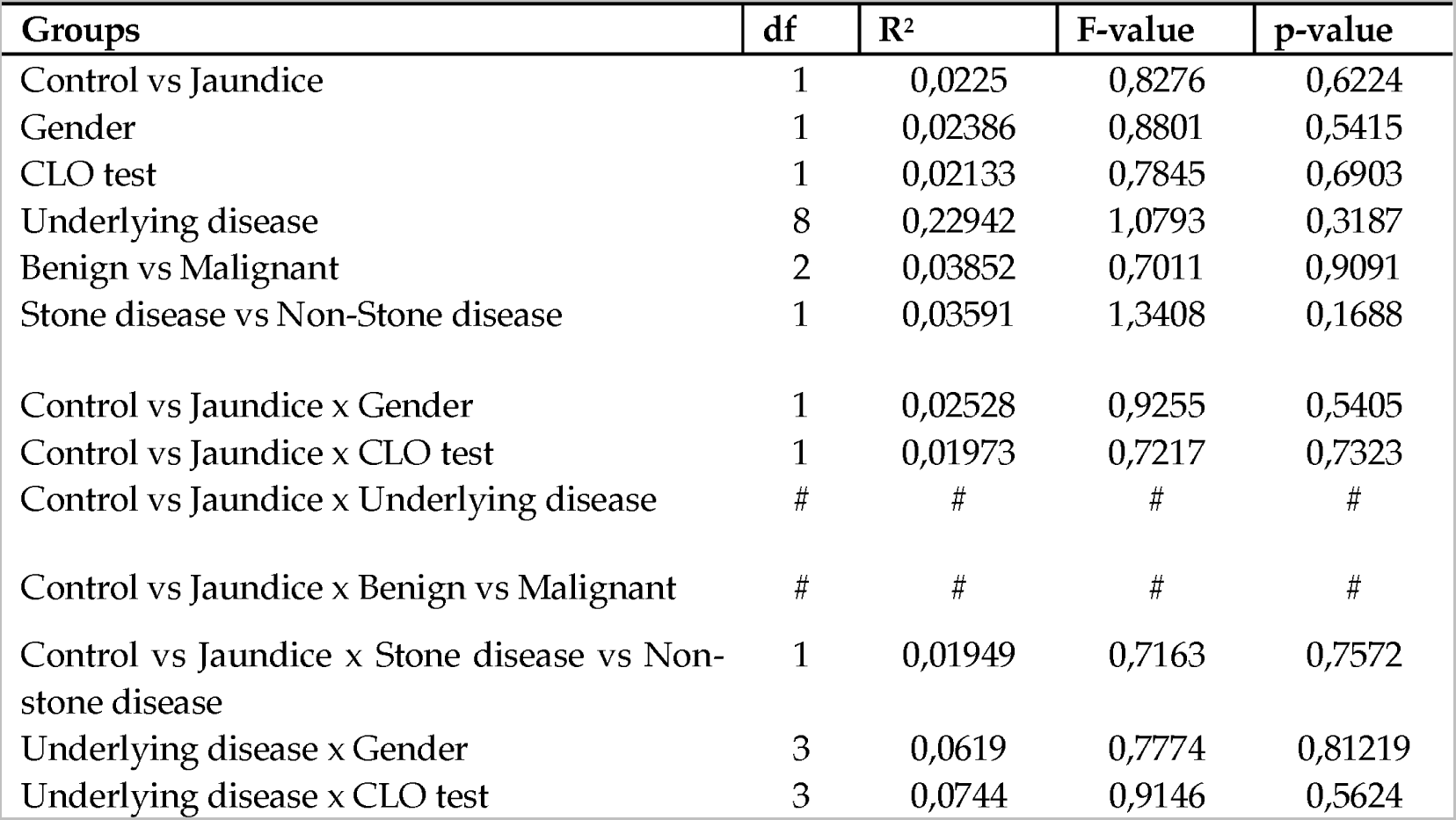
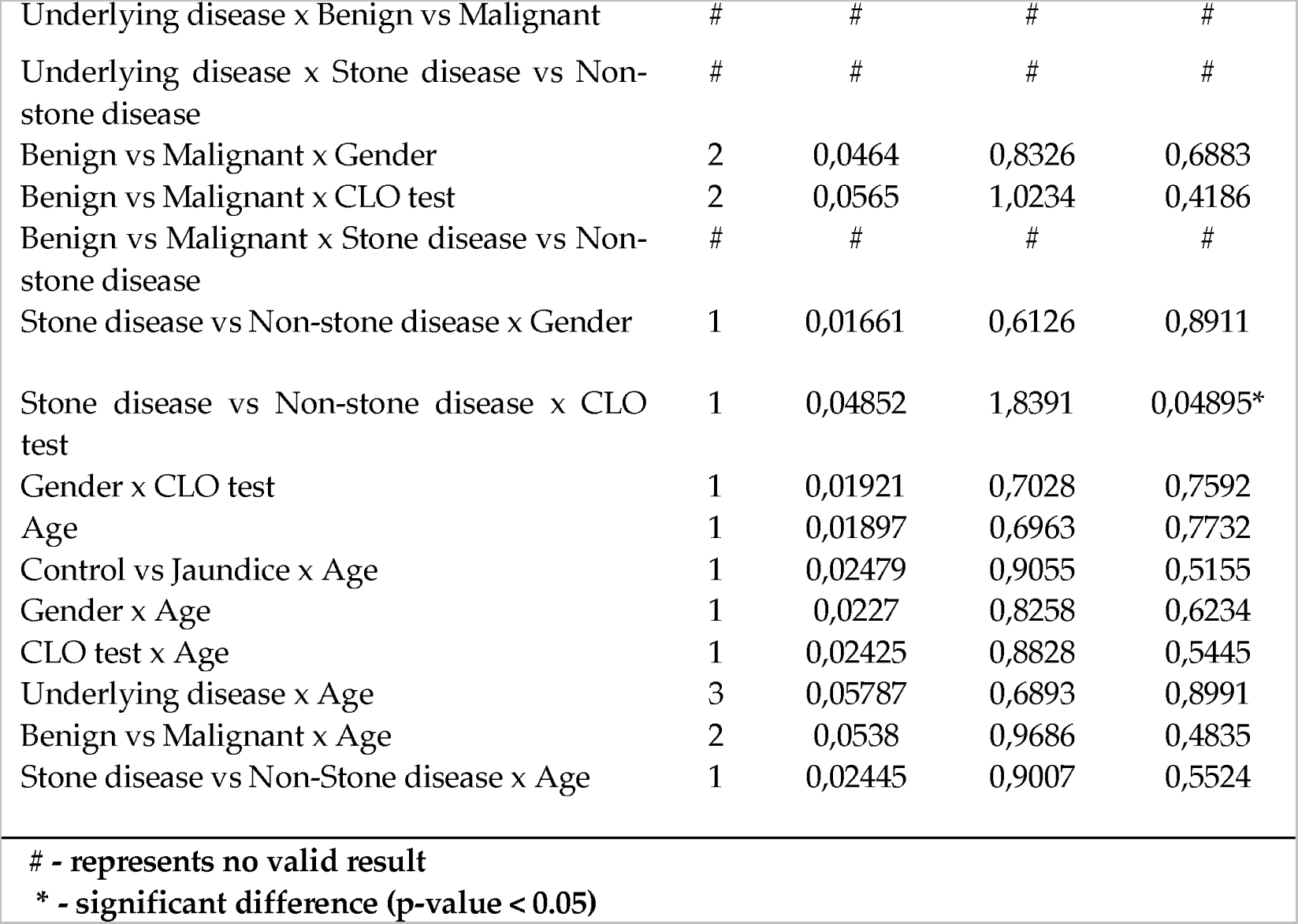
Bray-Curtis measure of dissimilarity of the beta-diversity of the microbiota of the duodenal mucosa factoring in multiple variables.

**Table S7:**
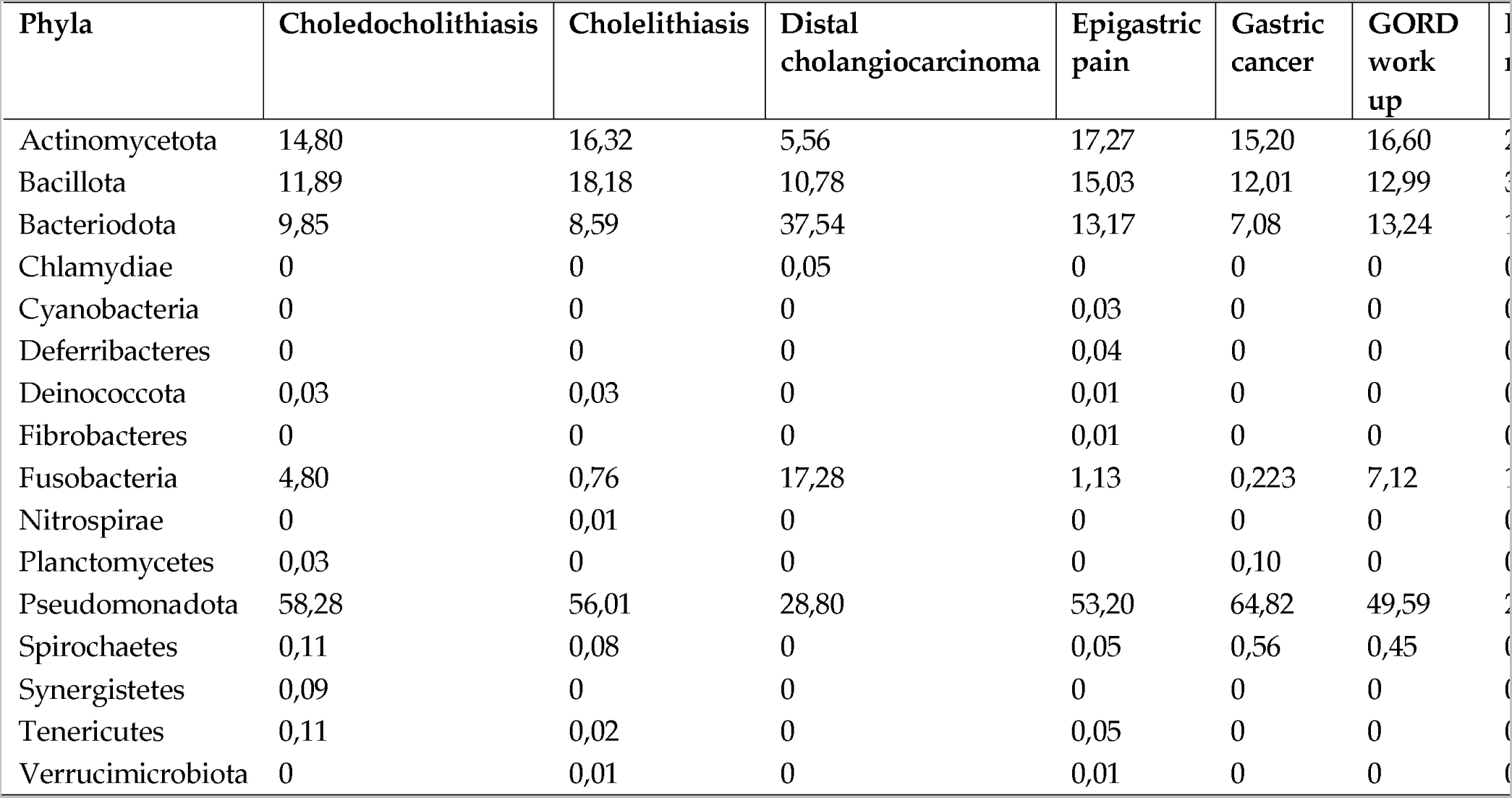
Relative abundances, expressed as a percentage, of the bacterial phyla in the duodenum of patients, comparing patients based on the underlying conditions at the time of endoscopy

| Phyla | Choledocholithiasis | Cholelithiasis | Distal cholangiocarcinoma | Epigastric pain | Gastric cancer | GORD work up |  |
| --- | --- | --- | --- | --- | --- | --- | --- |
| Actinomycetota | 14,80 | 16,32 | 5,56 | 17,27 | 15,20 | 16,60 | 2 |
| Bacillota | 11,89 | 18,18 | 10,78 | 15,03 | 12,01 | 12,99 | 3 |
| Bacteriodota | 9,85 | 8,59 | 37,54 | 13,17 | 7,08 | 13,24 | 1 |
| Chlamydiae | 0 | 0 | 0,05 | 0 | 0 | 0 | 0 |
| Cyanobacteria | 0 | 0 | 0 | 0,03 | 0 | 0 | 0 |
| Deferribacteres | 0 | 0 | 0 | 0,04 | 0 | 0 | 0 |
| Deinococcota | 0,03 | 0,03 | 0 | 0,01 | 0 | 0 | 0 |
| Fibrobacteres | 0 | 0 | 0 | 0,01 | 0 | 0 | 0 |
| Fusobacteria | 4,80 | 0,76 | 17,28 | 1,13 | 0,223 | 7,12 | 1 |
| Nitrospirae | 0 | 0,01 | 0 | 0 | 0 | 0 | 0 |
| Planctomycetes | 0,03 | 0 | 0 | 0 | 0,10 | 0 | 0 |
| Pseudomonadota | 58,28 | 56,01 | 28,80 | 53,20 | 64,82 | 49,59 | 2 |
| Spirochaetes | 0,11 | 0,08 | 0 | 0,05 | 0,56 | 0,45 | 0 |
| Synergistetes | 0,09 | 0 | 0 | 0 | 0 | 0 | 0 |
| Tenericutes | 0,11 | 0,02 | 0 | 0,05 | 0 | 0 | 0 |
| Verrucimicrobiota | 0 | 0,01 | 0 | 0,01 | 0 | 0 | 0 |

**Table S8:**
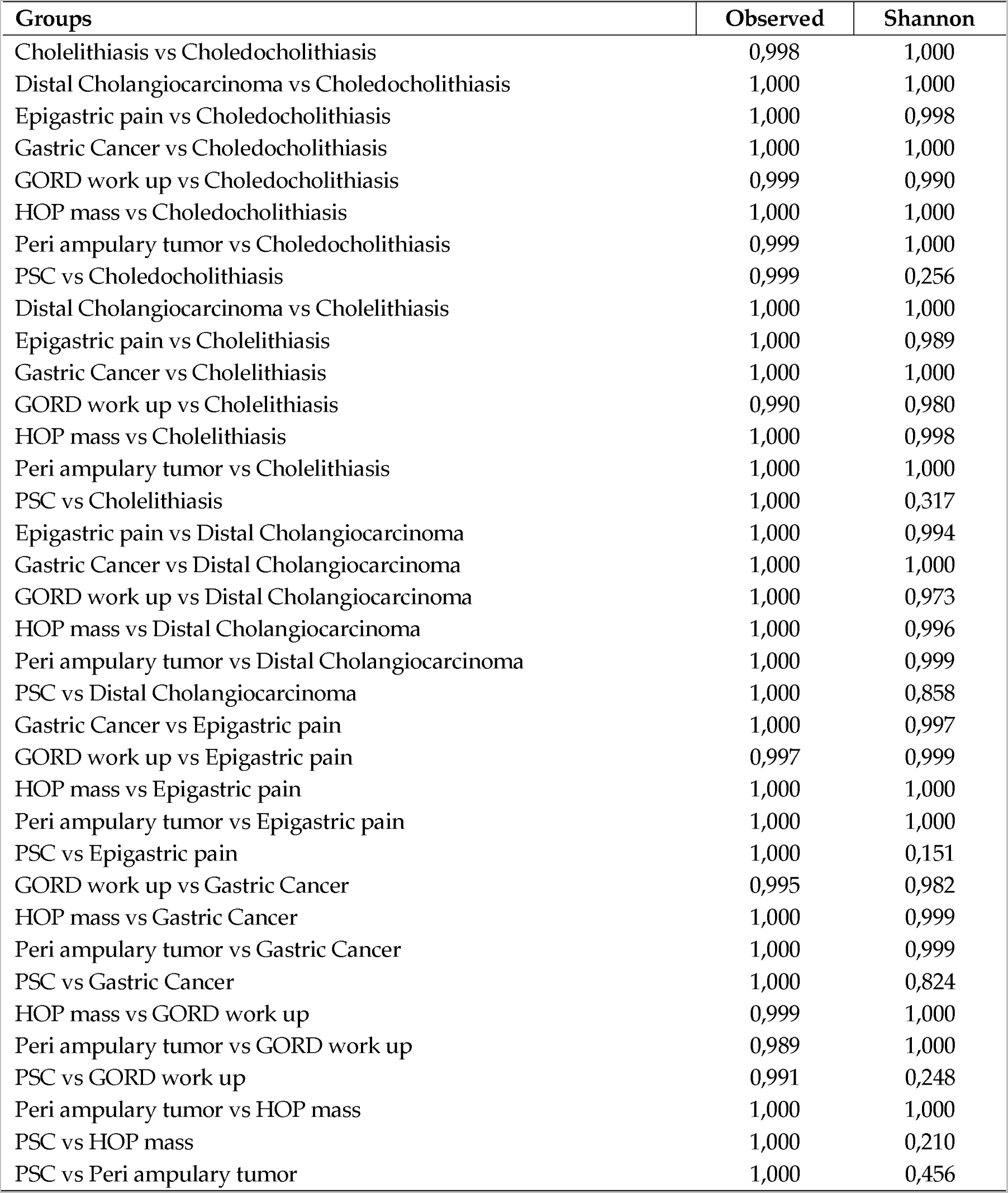
Impact of the variables on the bacterial alpha diversity indices computed using Tukey HSD test.

**Table S9:**
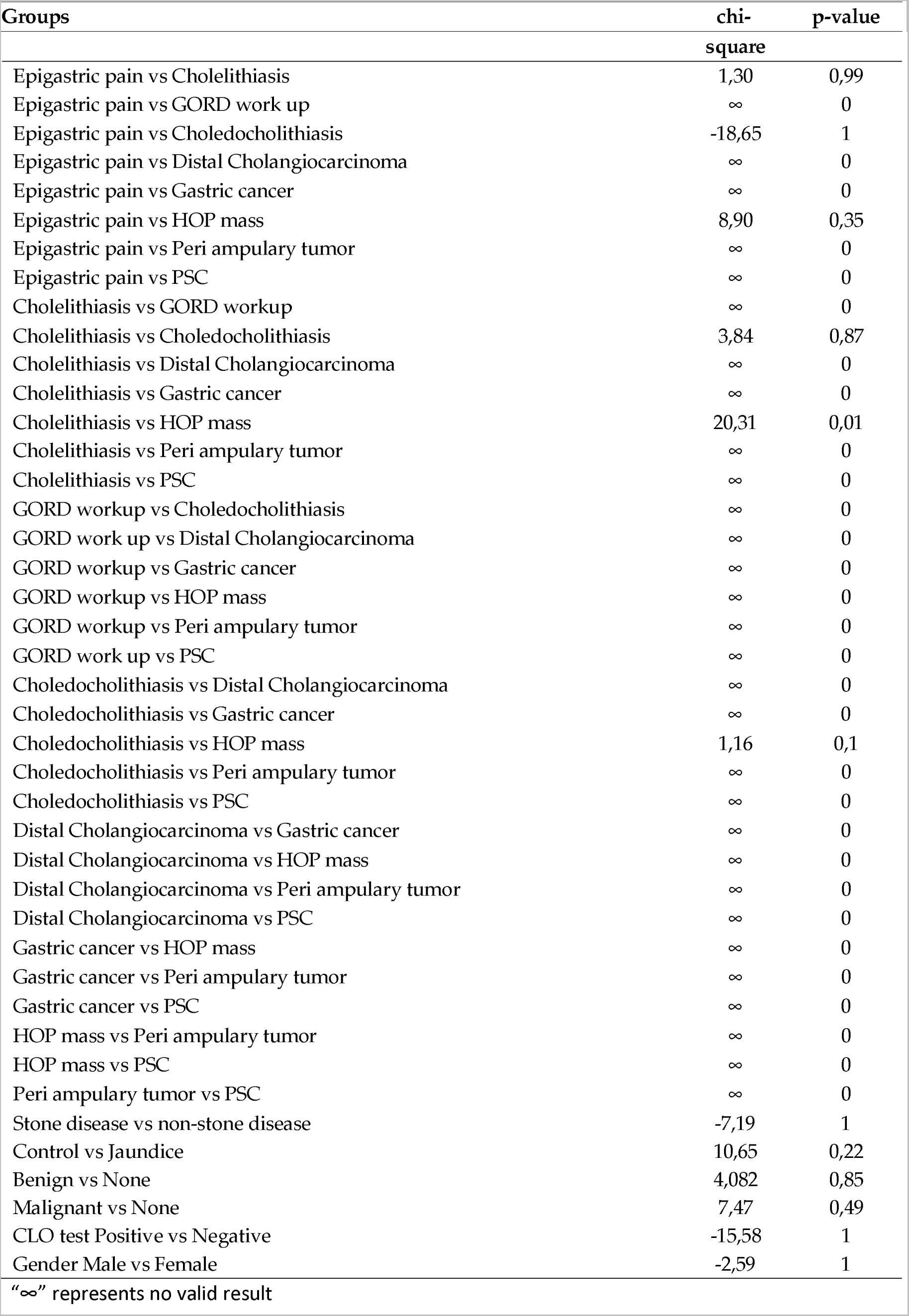
Impact of the multiple variables on the bacterial alpha diversity indices within the duodenal mucosa samples obtained by endoscopic biospies, these differences were calculated using Dirichlet-Multinomial distribution test.

