## Supplementary material for "Metataxonomic analysis demonstrates a shift in duodenal microbiota in South African patients with obstructive jaundice: A pilot study": Supplentary tables

**SUPPLEMENTARY TABLES**

Table S1: Diversity indices of individual samples in the jaundice group

|  | Observed | Chao1 | se,chao1 | ACE | se,ACE | Shannon | Simpson | | InvSimpson | Fisher | |
| --- | --- | --- | --- | --- | --- | --- | --- | --- | --- | --- | --- |
| S020 | 75,00 | 75,00 | 0,00 | 75,00 | 2,81 | 2,13 | 0,75 | 4,02 | | | 8,84 |
| S021 | 51,00 | 51,00 | 0,00 | NA | NA | 3,04 | 0,91 | 11,44 | | | 5,72 |
| S022 | 35,00 | 35,00 | 0,00 | NA | NA | 2,63 | 0,88 | 8,40 | | | 3,75 |
| S023 | 113,00 | 116,00 | 4,18 | 113,94 | 5,13 | 2,74 | 0,89 | 8,73 | | | 14,10 |
| S024 | 62,00 | 62,00 | 0,00 | 62,00 | 0,99 | 2,57 | 0,80 | 4,96 | | | 7,13 |
| S025 | 77,00 | 77,00 | 0,50 | 77,55 | 1,46 | 3,05 | 0,92 | 12,88 | | | 9,11 |
| S026 | 61,00 | 61,00 | 0,00 | 61,00 | 1,69 | 2,96 | 0,90 | 10,49 | | | 7,00 |
| S027 | 87,00 | 87,00 | 0,50 | 87,16 | 2,65 | 3,34 | 0,94 | 16,60 | | | 10,46 |
| S028 | 87,00 | 87,00 | 0,00 | 87,00 | 2,84 | 2,48 | 0,75 | 4,02 | | | 10,46 |
| S029 | 60,00 | 60,00 | 0,25 | 60,23 | 3,25 | 1,47 | 0,52 | 2,10 | | | 6,87 |
| S030 | 73,00 | 74,00 | 2,33 | 74,00 | 2,34 | 2,49 | 0,82 | 5,57 | | | 8,57 |
| S031 | 49,00 | 49,00 | 0,00 | 49,00 | 0,99 | 2,92 | 0,91 | 10,57 | | | 5,47 |
| S032 | 65,00 | 65,00 | 0,00 | 65,00 | 0,99 | 2,66 | 0,82 | 5,65 | | | 7,52 |
| S033 | 77,00 | 77,00 | 0,00 | NA | NA | 3,37 | 0,94 | 16,87 | | | 9,11 |
| S034 | 62,00 | 62,00 | 0,00 | NA | NA | 2,73 | 0,83 | 5,75 | | | 7,13 |
| S035 | 86,00 | 86,00 | 0,00 | NA | NA | 3,52 | 0,95 | 19,03 | | | 10,33 |
| S036 | 79,00 | 79,00 | 0,00 | 79,00 | 2,16 | 2,23 | 0,80 | 4,91 | | | 9,38 |
| S037 | 61,00 | 61,00 | 0,00 | 61,00 | 0,99 | 3,13 | 0,92 | 13,12 | | | 7,00 |
| S038 | 78,00 | 78,00 | 0,00 | 78,00 | 1,40 | 2,32 | 0,74 | 3,82 | | | 9,24 |

Table S2: Mean distribution of phyla comparing control and jaundice cohorts

| **Phyla** | **Control** | **Jaundice** |
| --- | --- | --- |
| *Arthrobacter* | 6,07 | 4,94 |
| *Fusobacterium* | 1,19 | 4,03 |
| *Haemophilus* | 4,00 | 2,39 |
| *Helicobacter* | 6,23 | 2,36 |
| *Pelomonas* | 8,29 | 6,43 |
| *Prevotella* | 7,42 | 9,55 |
| *Propionibacterium* | 3,69 | 5,48 |
| *Pseudomonas* | 11,02 | 14,59 |
| *Ralstonia* | 3,31 | 2,13 |
| *Streptococcus* | 6,53 | 5,20 |
| *Veillonella* | 3,62 | 3,72 |
| Other | 38,64 | 39,19 |

Table S3: Mean distribution of genera comparing patients with cholelithiasis and HOP (head of pancreas) mass

| **Genera** | **Cholelithiasis** | **HOP mass** |
| --- | --- | --- |
| *Arthrobacter* | 5,45 | 4,40 |
| *Bergeyella* | 0 | 3,63 |
| *Haemophilus* | 4,09 | 1,03 |
| *Helicobacter* | 4,42 | 0,03 |
| *Microbacterium* | 1,38 | 3,78 |
| *Pelomonas* | 7,92 | 6,94 |
| *Prevotella* | 5,46 | 4,45 |
| *Propionibacterium* | 3,43 | 7,24 |
| *Pseudomonas* | 21,14 | 4,66 |
| *Ralstonia* | 2,69 | 1,91 |
| *Staphylococcus* | 0,78 | 3,21 |
| *Streptococcus* | 7,81 | 10,80 |
| *Veillonella* | 5,22 | 12,46 |
| *Other* | 23,43 | 26,04 |

Table S4: Relative abundances, expressed as a percentage, of the bacterial phyla in the duodenum of patients when grouped according to the presence or absence of stone disease.

| **Phyla** | **Stone disease** | **Non-stone disease** |
| --- | --- | --- |
| Actinobacteria | 14,90 | 18,44 |
| Bacillota | 15,13 | 18,71 |
| Bacteriodota | 12,11 | 13,54 |
| Chlamydiae | 0 | 0 |
| Cyanobacteria | 0 | 0,02 |
| Deferribacteres | 0 | 0,02 |
| Deinococcota | 0,03 | 0,01 |
| Fibrobacteres | 0 | 0 |
| Fusobacteria | 2,89 | 2,43 |
| Nitrospirae | 0 | 0 |
| Planctomycetes | 0,02 | 0,01 |
| Pseudomonadota | 54,62 | 46,54 |
| Spirochaetes | 0,18 | 0,13 |
| Synergistetes | 0,05 | 0 |
| Tenericutes | 0,07 | 0,13 |
| Verrucimicrobiota | 0 | 0,01 |

Table S5: Relative abundances, expressed as a percentage, of the bacterial phyla in the duodenum of patients, comparing patients with benign and malignant conditions for jaundiced as well as control patients

| **Phyla** | **Control** | **Malignant** | **Benign** |
| --- | --- | --- | --- |
| Actinomycetota | 16,79 | 20,21 | 13,83 |
| Bacillota | 16,41 | 24,25 | 12,84 |
| Bacteriodota | 11,00 | 14,07 | 14,76 |
| Chlamydiae | 0 | 0,01 | 0 |
| Cyanobacteria | 0,02 | 0 | 0 |
| Deferribacteres | 0,02 | 0 | 0 |
| Deinococcota | 0,02 | 0,01 | 0,03 |
| Fibrobacteres | 0,01 | 0 | 0 |
| Fusobacteria | 1,27 | 3,44 | 4,49 |
| Nitrospirae | 0 | 0 | 0 |
| Planctomycetes | 0 | 0,01 | 0,03 |
| Proteobacteria | 54,34 | 37,56 | 53,59 |
| Spirochaetes | 0,08 | 0,18 | 0,25 |
| Synergistetes | 0 | 0,003764 | 0,08 |
| Tenericutes | 0,03 | 0,260849 | 0,10 |
| Verrucimicrobiota | 0,01 | 0 | 0 |

Table S6: Bray-Curtis measure of dissimilarity of the beta-diversity of the microbiota of the duodenal mucosa factoring in multiple variables.

| **Groups** | **df** | **R^2^** | **F-value** | **p-value** |
| --- | --- | --- | --- | --- |
| Control vs Jaundice | 1 | 0,0225 | 0,8276 | 0,6224 |
| Gender | 1 | 0,02386 | 0,8801 | 0,5415 |
| CLO test | 1 | 0,02133 | 0,7845 | 0,6903 |
| Underlying disease | 8 | 0,22942 | 1,0793 | 0,3187 |
| Benign vs Malignant | 2 | 0,03852 | 0,7011 | 0,9091 |
| Stone disease vs Non-Stone disease | 1 | 0,03591 | 1,3408 | 0,1688 |
| Control vs Jaundice x Gender | 1 | 0,02528 | 0,9255 | 0,5405 |
| Control vs Jaundice x CLO test | 1 | 0,01973 | 0,7217 | 0,7323 |
| Control vs Jaundice x Underlying disease | # | # | # | # |
| Control vs Jaundice x Benign vs Malignant | # | # | # | # |
| Control vs Jaundice x Stone disease vs Non-stone disease | 1 | 0,01949 | 0,7163 | 0,7572 |
| Underlying disease x Gender | 3 | 0,0619 | 0,7774 | 0,81219 |
| Underlying disease x CLO test | 3 | 0,0744 | 0,9146 | 0,5624 |
| Underlying disease x Benign vs Malignant | # | # | # | # |
| Underlying disease x Stone disease vs Non-stone disease | # | # | # | # |
| Benign vs Malignant x Gender | 2 | 0,0464 | 0,8326 | 0,6883 |
| Benign vs Malignant x CLO test | 2 | 0,0565 | 1,0234 | 0,4186 |
| Benign vs Malignant x Stone disease vs Non-stone disease | # | # | # | # |
| Stone disease vs Non-stone disease x Gender | 1 | 0,01661 | 0,6126 | 0,8911 |
| Stone disease vs Non-stone disease x CLO test | 1 | 0,04852 | 1,8391 | 0,04895* |
| Gender x CLO test | 1 | 0,01921 | 0,7028 | 0,7592 |
| Age | 1 | 0,01897 | 0,6963 | 0,7732 |
| Control vs Jaundice x Age | 1 | 0,02479 | 0,9055 | 0,5155 |
| Gender x Age | 1 | 0,0227 | 0,8258 | 0,6234 |
| CLO test x Age | 1 | 0,02425 | 0,8828 | 0,5445 |
| Underlying disease x Age | 3 | 0,05787 | 0,6893 | 0,8991 |
| Benign vs Malignant x Age | 2 | 0,0538 | 0,9686 | 0,4835 |
| Stone disease vs Non-Stone disease x Age | 1 | 0,02445 | 0,9007 | 0,5524 |

\# - represents no valid result

* - significant difference (p-value < 0.05)

Table S7: Relative abundances, expressed as a percentage, of the bacterial phyla in the duodenum of patients, comparing patients based on the underlying conditions at the time of endoscopy

| **Phyla** | **Choledocholithiasis** | **Cholelithiasis** | **Distal cholangiocarcinoma** | **Epigastric pain** | **Gastric cancer** | **GORD work up** | **HOP mass** | **Peri ampullary tumor** | **PSC** |
| --- | --- | --- | --- | --- | --- | --- | --- | --- | --- |
| Actinomycetota | 14,80 | 16,32 | 5,56 | 17,27 | 15,20 | 16,60 | 21,52 | 34,61 | 3,14 |
| Bacillota | 11,89 | 18,18 | 10,78 | 15,03 | 12,01 | 12,99 | 34,19 | 10,23 | 23,36 |
| Bacteriodota | 9,85 | 8,59 | 37,54 | 13,17 | 7,08 | 13,24 | 12,11 | 5,39 | 68,71 |
| Chlamydiae | 0 | 0 | 0,05 | 0 | 0 | 0 | 0 | 0 | 0 |
| Cyanobacteria | 0 | 0 | 0 | 0,03 | 0 | 0 | 0 | 0 | 0 |
| Deferribacteres | 0 | 0 | 0 | 0,04 | 0 | 0 | 0 | 0 | 0 |
| Deinococcota | 0,03 | 0,03 | 0 | 0,01 | 0 | 0 | 0,02 | 0,013 | 0 |
| Fibrobacteres | 0 | 0 | 0 | 0,01 | 0 | 0 | 0 | 0 | 0 |
| Fusobacteria | 4,80 | 0,76 | 17,28 | 1,13 | 0,223 | 7,12 | 1,52 | 0,49 | 1,02 |
| Nitrospirae | 0 | 0,01 | 0 | 0 | 0 | 0 | 0,01 | 0 | 0 |
| Planctomycetes | 0,03 | 0 | 0 | 0 | 0,10 | 0 | 0 | 0 | 0 |
| Pseudomonadota | 58,28 | 56,01 | 28,80 | 53,20 | 64,82 | 49,59 | 29,99 | 49,26 | 1,93 |
| Spirochaetes | 0,11 | 0,08 | 0 | 0,05 | 0,56 | 0,45 | 0,17 | 0 | 1,84 |
| Synergistetes | 0,09 | 0 | 0 | 0 | 0 | 0 | 0,01 | 0 | 0 |
| Tenericutes | 0,11 | 0,02 | 0 | 0,05 | 0 | 0 | 0,46 | 0 | 0 |
| Verrucimicrobiota | 0 | 0,01 | 0 | 0,01 | 0 | 0 | 0 | 0 | 0 |

Table S8: Impact of the multiple variables on the bacterial alpha diversity indices computed using Tukey HSD test.

| **Groups** | **Observed** | **Shannon** |
| --- | --- | --- |
| Cholelithiasis vs Choledocholithiasis | 0,998 | 1,000 |
| Distal Cholangiocarcinoma vs Choledocholithiasis | 1,000 | 1,000 |
| Epigastric pain vs Choledocholithiasis | 1,000 | 0,998 |
| Gastric Cancer vs Choledocholithiasis | 1,000 | 1,000 |
| GORD work up vs Choledocholithiasis | 0,999 | 0,990 |
| HOP mass vs Choledocholithiasis | 1,000 | 1,000 |
| Peri ampulary tumor vs Choledocholithiasis | 0,999 | 1,000 |
| PSC vs Choledocholithiasis | 0,999 | 0,256 |
| Distal Cholangiocarcinoma vs Cholelithiasis | 1,000 | 1,000 |
| Epigastric pain vs Cholelithiasis | 1,000 | 0,989 |
| Gastric Cancer vs Cholelithiasis | 1,000 | 1,000 |
| GORD work up vs Cholelithiasis | 0,990 | 0,980 |
| HOP mass vs Cholelithiasis | 1,000 | 0,998 |
| Peri ampulary tumor vs Cholelithiasis | 1,000 | 1,000 |
| PSC vs Cholelithiasis | 1,000 | 0,317 |
| Epigastric pain vs Distal Cholangiocarcinoma | 1,000 | 0,994 |
| Gastric Cancer vs Distal Cholangiocarcinoma | 1,000 | 1,000 |
| GORD work up vs Distal Cholangiocarcinoma | 1,000 | 0,973 |
| HOP mass vs Distal Cholangiocarcinoma | 1,000 | 0,996 |
| Peri ampulary tumor vs Distal Cholangiocarcinoma | 1,000 | 0,999 |
| PSC vs Distal Cholangiocarcinoma | 1,000 | 0,858 |
| Gastric Cancer vs Epigastric pain | 1,000 | 0,997 |
| GORD work up vs Epigastric pain | 0,997 | 0,999 |
| HOP mass vs Epigastric pain | 1,000 | 1,000 |
| Peri ampulary tumor vs Epigastric pain | 1,000 | 1,000 |
| PSC vs Epigastric pain | 1,000 | 0,151 |
| GORD work up vs Gastric Cancer | 0,995 | 0,982 |
| HOP mass vs Gastric Cancer | 1,000 | 0,999 |
| Peri ampulary tumor vs Gastric Cancer | 1,000 | 0,999 |
| PSC vs Gastric Cancer | 1,000 | 0,824 |
| HOP mass vs GORD work up | 0,999 | 1,000 |
| Peri ampulary tumor vs GORD work up | 0,989 | 1,000 |
| PSC vs GORD work up | 0,991 | 0,248 |
| Peri ampulary tumor vs HOP mass | 1,000 | 1,000 |
| PSC vs HOP mass | 1,000 | 0,210 |
| PSC vs Peri ampulary tumor | 1,000 | 0,456 |

Table S9: Impact of the multiple variables on the bacterial alpha diversity indices within the duodenal mucosa samples obtained by endoscopic biospies, these differences were calculated using Dirichlet-Multinomial distribution test.

| **Groups** | **chi-square** | **p-value** |
| --- | --- | --- |
| Epigastric pain vs Cholelithiasis | 1,30 | 0,99 |
| Epigastric pain vs GORD work up | ∞ | 0 |
| Epigastric pain vs Choledocholithiasis | -18,65 | 1 |
| Epigastric pain vs Distal Cholangiocarcinoma | ∞ | 0 |
| Epigastric pain vs Gastric cancer | ∞ | 0 |
| Epigastric pain vs HOP mass | 8,90 | 0,35 |
| Epigastric pain vs Peri ampulary tumor | ∞ | 0 |
| Epigastric pain vs PSC | ∞ | 0 |
| Cholelithiasis vs GORD workup | ∞ | 0 |
| Cholelithiasis vs Choledocholithiasis | 3,84 | 0,87 |
| Cholelithiasis vs Distal Cholangiocarcinoma | ∞ | 0 |
| Cholelithiasis vs Gastric cancer | ∞ | 0 |
| Cholelithiasis vs HOP mass | 20,31 | 0,01 |
| Cholelithiasis vs Peri ampulary tumor | ∞ | 0 |
| Cholelithiasis vs PSC | ∞ | 0 |
| GORD workup vs Choledocholithiasis | ∞ | 0 |
| GORD work up vs Distal Cholangiocarcinoma | ∞ | 0 |
| GORD workup vs Gastric cancer | ∞ | 0 |
| GORD workup vs HOP mass | ∞ | 0 |
| GORD workup vs Peri ampulary tumor | ∞ | 0 |
| GORD work up vs PSC | ∞ | 0 |
| Choledocholithiasis vs Distal Cholangiocarcinoma | ∞ | 0 |
| Choledocholithiasis vs Gastric cancer | ∞ | 0 |
| Choledocholithiasis vs HOP mass | 1,16 | 0,1 |
| Choledocholithiasis vs Peri ampulary tumor | ∞ | 0 |
| Choledocholithiasis vs PSC | ∞ | 0 |
| Distal Cholangiocarcinoma vs Gastric cancer | ∞ | 0 |
| Distal Cholangiocarcinoma vs HOP mass | ∞ | 0 |
| Distal Cholangiocarcinoma vs Peri ampulary tumor | ∞ | 0 |
| Distal Cholangiocarcinoma vs PSC | ∞ | 0 |
| Gastric cancer vs HOP mass | ∞ | 0 |
| Gastric cancer vs Peri ampulary tumor | ∞ | 0 |
| Gastric cancer vs PSC | ∞ | 0 |
| HOP mass vs Peri ampulary tumor | ∞ | 0 |
| HOP mass vs PSC | ∞ | 0 |
| Peri ampulary tumor vs PSC | ∞ | 0 |
| Stone disease vs non-stone disease | -7,19 | 1 |
| Control vs Jaundice | 10,65 | 0,22 |
| Benign vs None | 4,082 | 0,85 |
| Malignant vs None | 7,47 | 0,49 |
| CLO test Positive vs Negative | -15,58 | 1 |
| Gender Male vs Female | -2,59 | 1 |

“∞” represents no valid result
