## Supplementary figures and images for "Metataxonomic analysis demonstrates a shift in duodenal microbiota in South African patients with obstructive jaundice: A pilot study"

### Figure S1

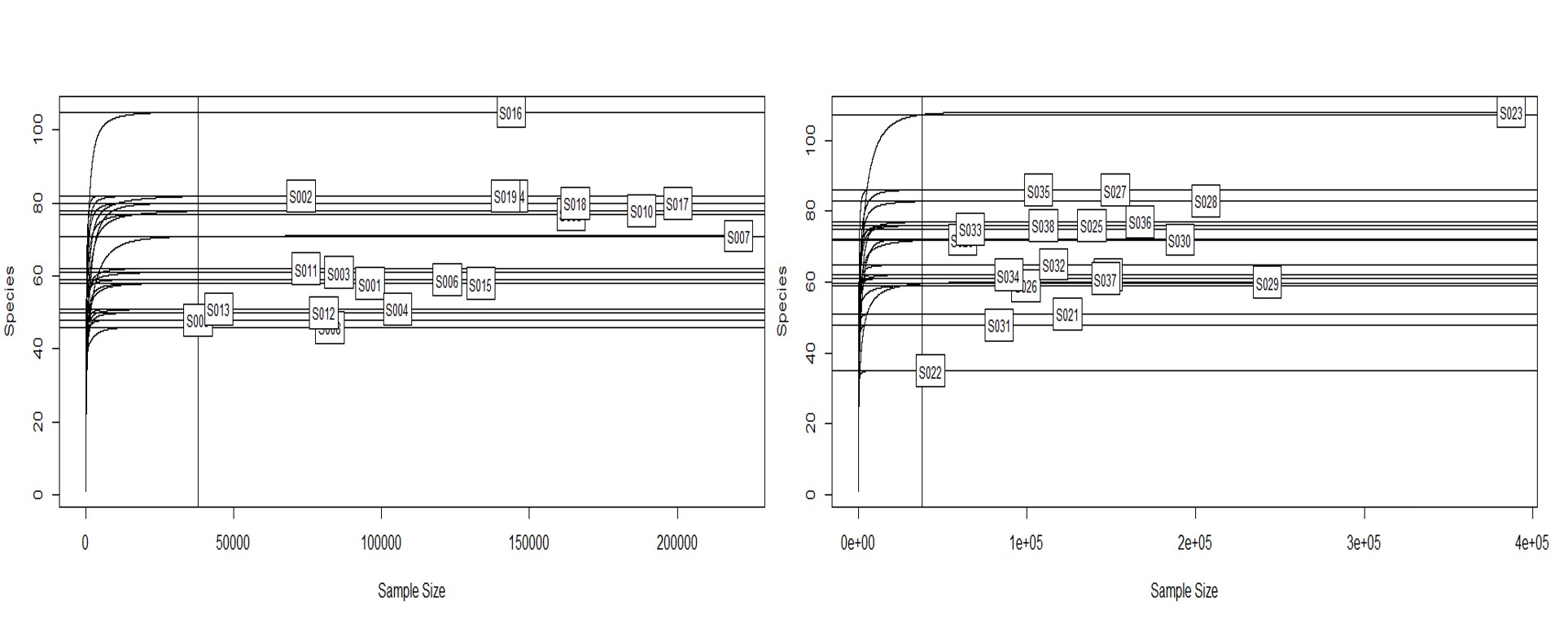

### Figure S2

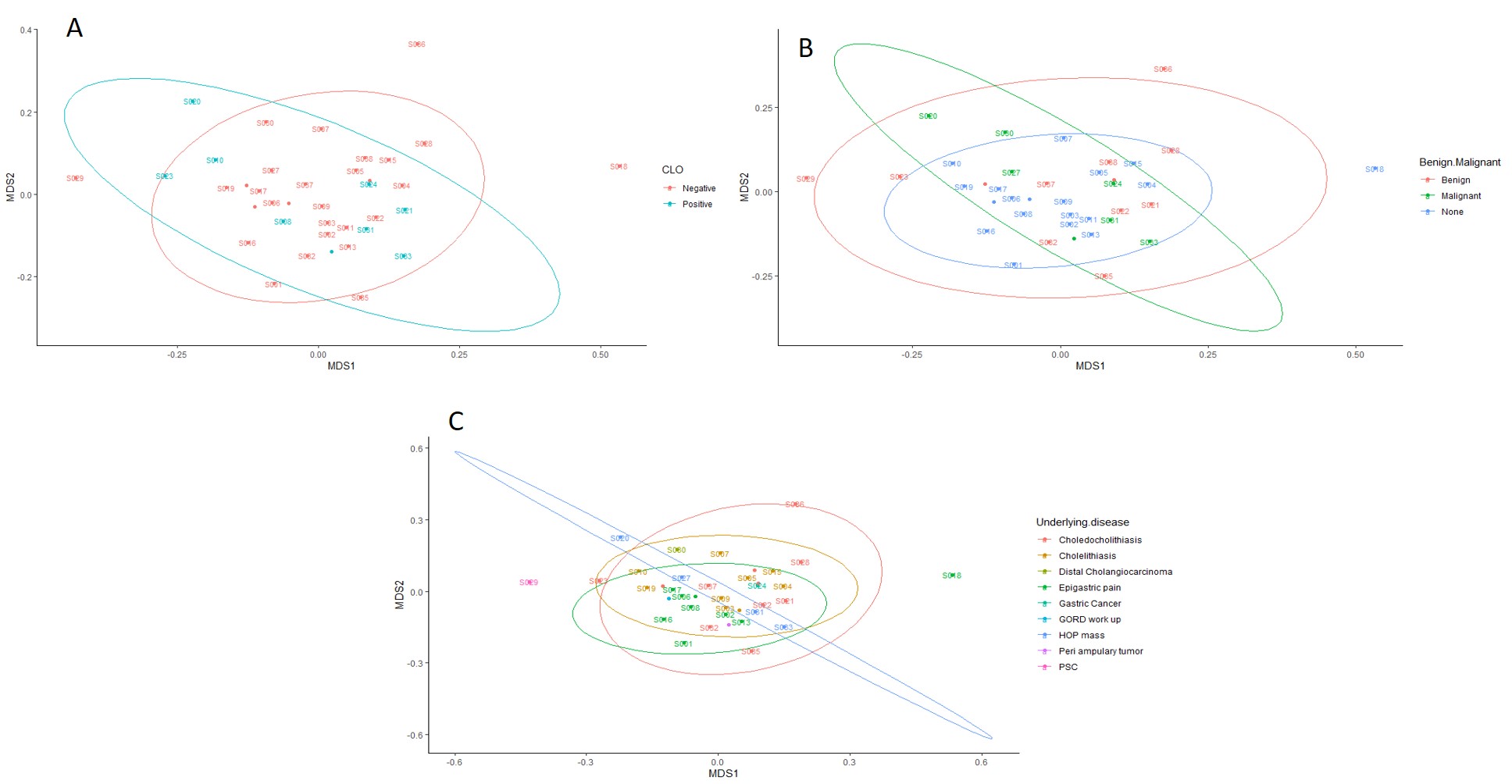
